# A novel mechanism of HIF2-dependent PLK1-mediated metastasis and drug resistance of clear cell Renal Cell Carcinoma

**DOI:** 10.1101/2020.02.05.20020552

**Authors:** Maeva Dufies, Annelies Verbiest, Lindsay S Cooley, Papa Diogop Ndiaye, Julien Viotti, Xingkang He, Nicolas Nottet, Wilfried Souleyreau, Anais Hagege, Stephanie Torrino, Julien Parola, Sandy Giuliano, Delphine Borchiellini, Renaud Schiappa, Baharia Mograbi, Jessica Zucman-Rossi, Karim Bensalah, Alain Ravaud, Patrick Auberger, Andréas Bikfalvi, Emmanuel Chamorey, Nathalie Rioux-Leclercq, Nathalie M. Mazure, Benoit Beuselinck, Yihai Cao, Jean Christophe Bernhard, Damien Ambrosetti, Gilles Pagès

## Abstract

Polo-Like Kinase 1 (Plk1) expression is inversely correlated with survival advantages in many cancers. However, molecular mechanisms that underlie Plk1 expression are poorly understood. Here, we uncover a novel hypoxia-regulated mechanism of Plk1-mediated cancer metastasis and drug resistance. We demonstrated that a new HIF-2-dependent regulatory pathway drives Plk1 expression in clear cell renal cell carcinoma (ccRCC). Mechanistically, HIF-2 transcriptionally targets the hypoxia response element of the Plk1 promoter. In ccRCC patients, high expression of Plk1 was correlated to poor disease-free survival and overall survival. Loss-of-function of Plk1 *in vivo* markedly attenuated ccRCC growth and metastasis. High Plk1 expression conferred a resistant phenotype of ccRCC to targeted therapeutics such as sunitinib, *in vitro, in vivo* and in metastatic ccRCC patients. Importantly, high Plk1 expression was defined in a subpopulation of ccRCC patients that are refractory to current therapies. Hence, we propose a therapeutic paradigm for improving outcomes of ccRCC patients.

## Introduction

The majority of ccRCC patients carry genetic aberrations of the *von Hippel-Lindau* (*VHL*) gene leading to genetic stabilisation of hypoxia-inducible factor (HIF) transcription factor. The HIF pathway drives tumor development and progression in the VHL–inactivated ccRCC. HIF transcriptionally targets over 100 genes (1), and the loss of VHL function induces constitutive HIF-1α/2α expression that markedly upregulated their targeted genes, including vascular endothelial growth factor (VEGF) and erythropoietin (EPO). Consequently, ccRCC is a hypervascularized tumor that carries frequent mutations in chromosome 3p, which affects an array of chromatin-remodelling genes, including *Polybromo 1* (*PBRM1*), *SET Domain Containing 2* (*SETD2*), and *BRCA1 Associated Protein 1* (*BAP1*) (2, 3). Tyrosine kinase inhibitors (TKI) primarily targeting VEGF receptors such as sunitinib are the first-line therapy for treating metastatic ccRCC (4). Immune checkpoint inhibitors have also approved as the first-line therapy in some countries. Sunitinib inhibits angiogenesis by blocking VEGFRs. Interestingly, it also directly inhibits ccRCC cell proliferation through non-VEGFR-mediated pathways. Nevertheless, clinical benefits are limited and transient in most cases and the majority of patients develop resistant over time (5, 6).

Based on gene expression, methylation status, mutation profile, cytogenetic anomalies, and immune cell infiltration, 4 subtypes of ccRCC patients (ccrcc1–4) have been classified (7-10). These markers have prognostic and predictive values for guiding TKI-based therapy. The ccrcc2&3 subtypes possess a good prognosis value of progression free (PFS) and overall survival (OS) and favourable for TKI therapy, whereas the ccrcc1&4 subtypes have the opposite prognostic values with poor prognosis and TKI responses. The ccrcc2-tumors often express proangiogenic genes and ccrcc3-tumors resembles gene expression profiling of healthy kidney tissue. Ccrcc4-tumors exhibit an immune-inflamed phenotype, but an exhausted tumor cell capacity by immune cells. The ccrcc1-tumors belong to an immune-cold phenotype almost without Lymphocyte infiltration (7-10). Therefore, the ccrcc2&3-tumors are favourable for TKI therapy and the ccrcc4-tumors are potentially beneficial responders to immunotherapy. In contrast, ccrcc1-tumors fail to respond to either therapies (7, 8). Sunitinib-resistant tumor cells acquire an enhanced ability to proliferate. Therefore, cell-cycle regulators may be perturbed in sunitinib-resistant ccRCC tumors. Polo-Like Kinase 1 (Plk1) is a serine/threonine kinase that acts during cell cycle progression (11). Plk1 inhibits p53, and p53 represses the Plk1 promoter (12). High Plk1 expression correlates with an advanced disease stage, histological grades, metastatic potentials, and short-term survival in various tumors (13, 14). The Plk1 inhibitor volasertib inhibits a variety of carcinoma cell lines and induces tumor regression in several experimental tumor models (15, 16).

In this study, we describe a novel molecular mechanism of the HIF-2-Plk1-mediated ccRCC metastasis and drug resistance. Plk1may also serve as prognostic marker to predict ccRCC progression and drug resistance. We propose a new theranostic paradigm by targeting Plk1 for treating sunitinib resistant ccRCC. We provide compelling experimental evidence to support our conclusions and relate our findings to clinical relevance.

## Results

### High levels of Plk1 mRNA correlate with the HIF pathway in various cancers

Plk1 expression correlated with shorter PFS and OS in cancers ((17), Supplementary Table S1). An *in-silico* analysis revealed the presence of a consensus HRE in the *Plk1* promoter (Supplementary Fig. S1A). Since HIF-1α and HIF-2α are regulated by protein stabilization, we investigated the correlation between Plk1 and mRNA levels of HIF-1/2α targets rather than with HIF-1α or HIF-2α mRNA levels (Ca9 (HIF-1α), Oct4 (HIF-2α) or Glut1 (HIF-1α and HIF-2α)) in the TCGA database (Supplementary Table S1). Plk1 expression correlated with HIF-1α and HIF-2α targets in breast, liver cancers and sarcoma, with HIF-1 targets in melanoma, two types of kidney, head and neck, lung, pancreatic cancers and with HIF-2 targets in ccRCC. Plk1 was independent of HIF-1α and HIF-2α in uterine cancers. Hence, Plk1 expression depends on HIF-1α and/or HIF-2α in most cancers including ccRCC.

### Plk1 is a marker of poor prognosis in ccRCC

Because of VHL inactivation, ccRCC represent a paradigm to assess the relationship between Plk1 and HIF-α. In TCGA Database, Plk1 mRNA levels correlated with disease free survival (DFS) and OS (Supplementary Fig. S2B and C), a correlation confirmed in an independent cohort of French patients (111 ccRCC M0; Supplementary Table S2A; Supplementary Fig. S3D and E). In our cohort, the Plk1 mRNA levels were higher in ccRCC as compared to healthy kidney (p<0.0001, Fig. 1A) and were increased in necrotic ccRCC (high level of hypoxia, p=0.0313, Supplementary Fig. S2D). Tumors with two inactivated *VHL* alleles presented higher Plk1 mRNA levels as compared to tumors with normal or with only one inactivated *VHL* allele (p=0.05, Fig. 1*B*). High levels of Plk1 mRNA correlated with shorter DFS (39.1 months *vs* > 100 months, p=0.0004, Fig. 1C) and OS (63 months *vs* > 100 months p=0.0005, Fig. 1D) in M0 patients. Plk1 mRNA levels represented an independent marker for DFS and OS of the Fuhrman grade in multivariate analyses (Fig. 1E and F). Plk1 mRNA levels were indicative of DFS for low grade tumors (Fuhrman 2; 48.2 months *vs* > 100 months, p= 0.0005, Fig. 1G) and high grade tumors (Fuhrman 3 and 4; 27.5 months *vs* > 100 months, p=0.0476, Fig. 1H) for M0 patients. Plk1 protein levels on tissue microarrays did not correlate with the Fuhrman grade or with the metastatic stage. High levels of Plk1 correlated with a shorter DFS (p=0.042) and OS (p=0.0243) in M0 patients and a shorter DFS/PFS (p= 0.0492) and OS (p= 0.0272) in M1 patients (Supplementary Table S2B and Supplementary Fig. S3). Plk1 is a prognostic marker of survival, independent of the metastatic status.

**Figure 1.**
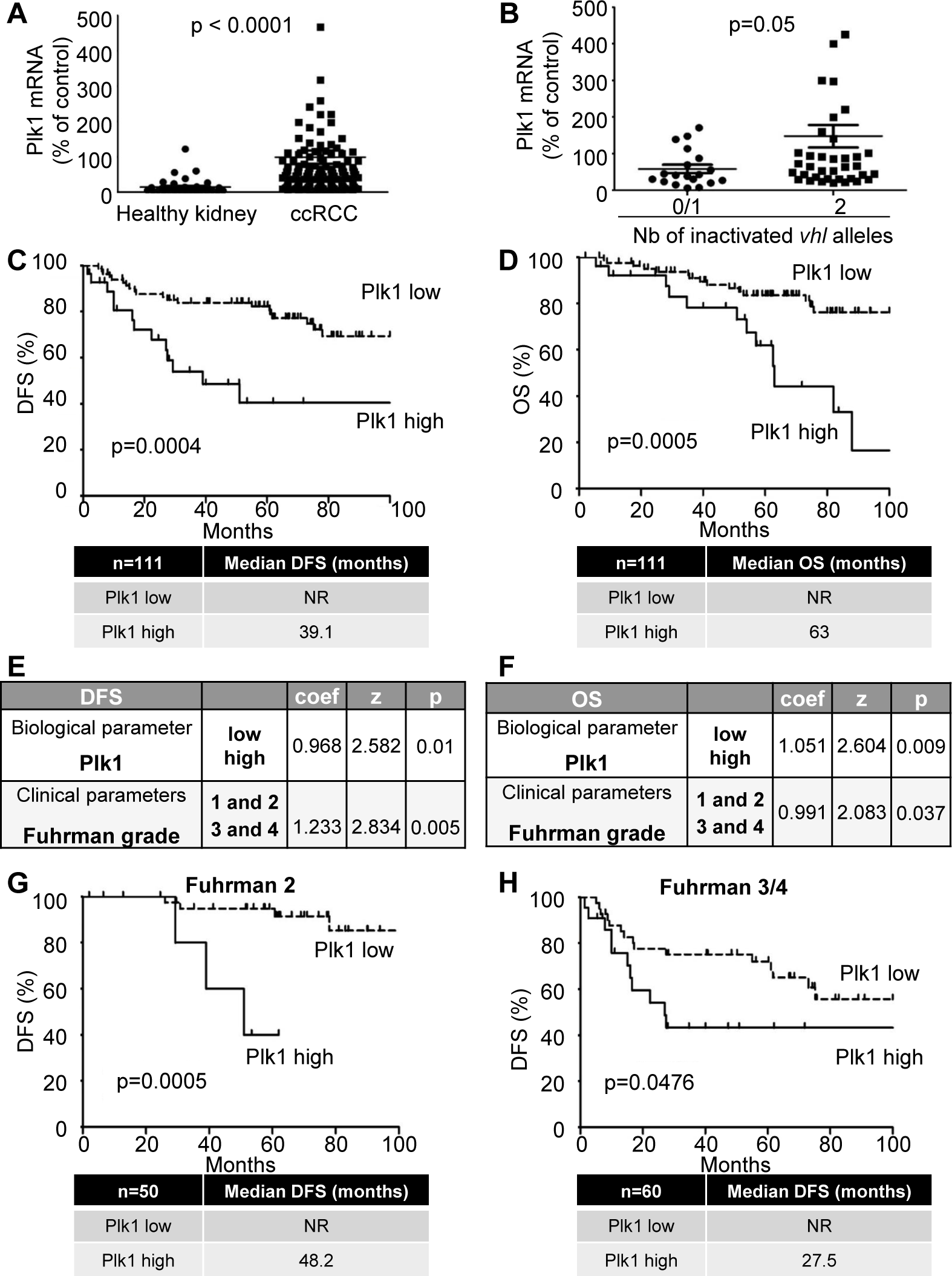
Plk1 is associated with poor prognosis in ccRCC. 111 ccRCC patients were analyzed for Plk1 mRNA levels in the kidneys. **A**, The levels of Plk1 mRNA in healthy kidney were compared with the levels in ccRCC. **B**, The levels of Plk1 mRNA in ccRCC patients with VHL-WT (0 or 1 inactivated vhl allele) were compared to the levels in ccRCC patients with VHL-inactivated (2 inactivated vhl alleles). **C and D**, The levels of Plk1 mRNA in 111 non-metastatic ccRCC patients correlated with DFS, **C**) or with OS, **D**). **E and F**, Multivariate analysis of Plk1, the Fuhrman grade and PFS (**E**) or OS (**F**). The multivariate analysis was performed using Cox regression adjusted to the Fuhrman grade. **G and H**, The levels of Plk1 mRNA in non-metastatic low grade (Fuhrman 2, **G**) or high grade (Fuhrman 3 and 4, **H**) ccRCC patients correlated with DFS. The third quartile value of Plk1 expression was chosen as the reference. For A and B, statistics were determined using an unpaired Student’s *t* test. For C, D, G and H, the Kaplan-Meier method was used to produce survival curves and analyses of censored data were performed using Cox models. Statistical significance (p values) is indicated. (see Supplementary Table S2A).

### HIF-2α binds to the *Plk1* promoter and stimulates its transcription in ccRCC cells

The relationship between hypoxia and Plk1 expression was further assessed in human ccRCC cell lines (RCC4 (R4), RCC10 (R10), 786-O (786), A498 (498), ACHN (A), Caki2 (C2) (Fig. 2A) and human primary normal (15S) and human primary ccRCC cells (TF, MM, CC, Fig. 2*B*) (18). HIF-1/2α were absent in cells with active VHL (A and C2, TF and 15S). Cells inactivated for VHL expressed HIF-1α and HIF-2α (R4, MM), or only HIF-2α (R10, 786, 498, CC). The VHL-active cell lines and primary tumor cells, presented low expression of Plk1 whereas VHL-inactivated cell lines and primary cells expressed Plk1 (Fig. 2A and B). Plk1 was absent in normal kidney cells (15S) that do not express HIF-1/2α. Chromatin immunoprecipitation (ChIP) analyses showed that only HIF-2α bound to the *Plk1* promoter (Fig. 2C). These results suggest direct regulation of *Plk1* transcription by HIF-2α but not by HIF-1α.

**Figure 2.**
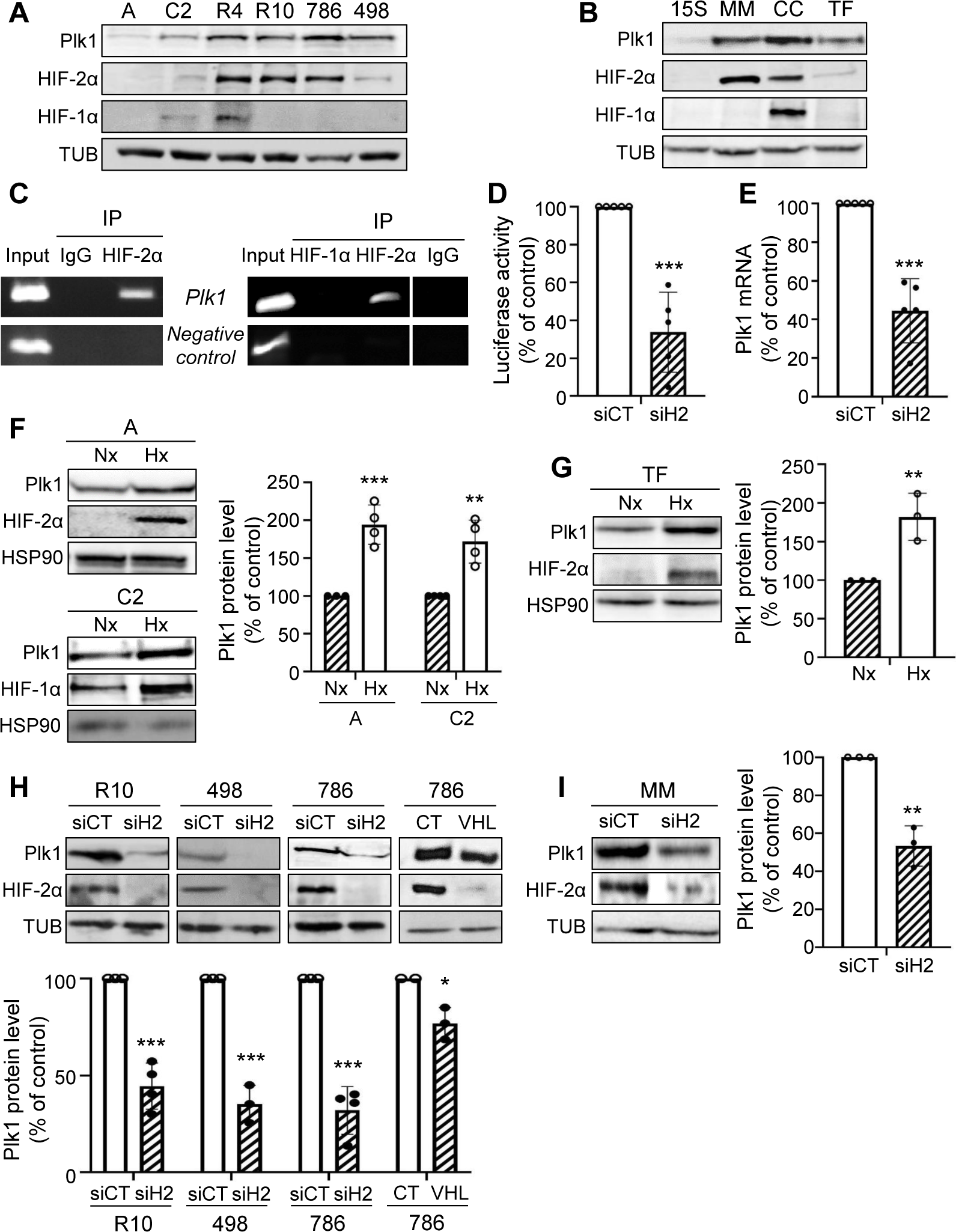
HIF2 bond to the *Plk1* promoter and regulated its expression in ccRCC cells. **A and B**, Different RCC cell lines [(ACHN (A), Caki2 (C2), RCC4 (R4), RCC10 (R10), 786-O (786) and A498 (498)] (**A**) or primary RCC cells (TF, MM and CC) and healthy renal cells (15S) (**B**) were evaluated for Plk1, HIF-1α and HIF-2α expression by immunoblotting. Tubulin (Tub) served as a loading control. **C**, ChIP experiments with HIF-2α and HIF-1α antibodies or negative CT antibodies were performed on extracts from 786 (right) and RCC4 (left) ccRCC cells. The promoter region of the *Plk1* promoter containing the HIF-α binding site was amplified by PCR. Results are representative of three independent experiments. **D**, ccRCC cell lines (VHL-inactivated) 786 were transfected with siRNA against HIF-2α (H2) for 24 h. Cells were then transfected with a renilla luciferase reporter gene under the control of the Plk1 promoter. The renilla luciferase activity normalized to the firefly luciferase (control vector) was the readout of the Plk1 promoter activity. **E**, 786 cells were transfected with siRNA against HIF-2α (H2) for 48 h. The Plk1 mRNA level was determined by qPCR. **F and G**, ccRCC cell lines (VHL-WT) A and C2 (**F**), or primary ccRCC cells (VHL-WT) TF (**G**) were cultured in normoxia (Nx) or hypoxia 1% O2 (Hx) for 24 h. Plk1, HIF-1α and HIF-2α expression were evaluated by immunoblotting. HSP90 served as a loading control. The graphs show the level of Plk1 (mean of three experiments). Control conditions were considered as the reference value (1). **H and I**, ccRCC cell lines (VHL-inactivated) R10, 498, 786 (**H**), or primary ccRCC cells (VHL-inactivated) MM (**I**) were transfected with siRNA against HIF-2α (H2) for 48 h. Plk1 and HIF-2α expression were evaluated by immunoblotting. HSP90 served as a loading control. The graphs show the level of Plk1. Control conditions were considered as the reference value (100). Results are represented by the means of three or more independent experiments (biological replication) ± SEM. Statistics were determined using an unpaired Student’s *t* test: * p<0.05, ** p<0.01, *** p<0.0001.

The role of HIF-α in *Plk1* transcription was evaluated by testing *Plk1* promoter activity and Plk1 mRNA levels after hypoxia or HIF-α down-regulation. HIF-2α-directed siRNA (siH2) decreased *Plk1* promoter activity (Fig. 2D) and Plk1 mRNA levels in VHL-inactivated ccRCC cell lines (Fig. 2E) and primary ccRCC cells only expressing HIF-2α (Supplementary Fig. S4A-D).

HIF-1α and HIF-2α down-regulation (siH1 and siH2) in a cell line (R4) and in primary ccRCC cells (CC) decreased the *Plk1* promoter activity, the amount of Plk1 mRNA and the level of Plk1 protein (Supplementary Fig. S4I and J). These results and the ChIP experiments suggested that HIF-1α indirectly regulates Plk1 expression. The Plk1 promoter activity, mRNA and protein amounts were very low in normal kidney cells (15S) and HIF-1α and HIF-2α down-regulation did not modify Plk1 levels (Supplementary Fig. S4I and J).

Following HIF-α stabilization by hypoxia in ccRCC cell lines or in primary ccRCC cells expressing active VHL, the promoter activity and Plk1 mRNA levels were up-regulated (Supplementary Fig. S4E-H).

Hypoxia stabilized HIF-1α in C2 and of HIF-2α in A and in TF primary cells resulting in Plk1 induction (Fig. 2F and G). siH2 inhibited Plk1 expression in VHL-inactivated ccRCC cell lines (R10, 498 and 786, Fig. 2H) and in MM primary ccRCC cells (Fig. 2I). Re-introduction of a functional VHL in 786 cells decreased Plk1 levels (Fig. 2H). Down-regulation of HIF-2α decreased Plk1 mRNA levels in R4 and CC cells (Supplementary Fig. S4K and L). These results suggest that *Plk1* is a HIF-2 target.

### SETD2 mutation stimulates Plk1 expression in ccRCC cells inactivated for VHL

ccRCC are frequently inactivated for VHL and mutations occur in chromatin-remodelling genes (PBRM1, BAP1 and SETD2). Mutations in PBRM1 and/or BAP1 did not modify Plk1 expression. However, tumors inactivated for VHL and SETD2, over-express Plk1 (Fig. 3A). Mutations in SETD2 in tumors with wild-type VHL (WT-VHL) did not over-express Plk1 (Fig. 3B). We therefore examined the mutational status of SETD2 in our ccRCC cell lines with inactivated or WT-VHL. 786, A, and C2 cells express normal SETD2 and 498 cells presented an inactivating mutation (*SETD2* V2536Efs*9). SETD2 down-regulation by siRNA in 498 cells did not modify Plk1 expression but SETD2 down-regulation in 786 cells increased mRNA and protein levels (Fig. 3C and E). The decrease in SETD2 in cells with active VHL did not alter Plk1 mRNA and protein levels (Fig. 3D and E). Our results suggested that inactivation of SETD2 modified Plk1 expression only when cells constitutively express HIF-2α. Therefore, we examined the link between SETD2 and HIF-2. siH2, or the expression of WT-VHL increased SETD2 mRNA and protein (Fig. 3G and I) levels. In contrast, hypoxia decreased SETD2 mRNA and protein (Fig. 3H and J) levels in WT-VHL cells. These results suggested that hypoxia stimulated Plk1 expression through downregulation of SETD2 leading to the accessibility of HIF-2 to the *Plk1* promoter and its subsequent transcriptional stimulation. Hence, an enhanced aggressiveness program involves Plk1 up-regulation through SETD2 inactivation and HIF-2α stabilization.

**Figure 3.**
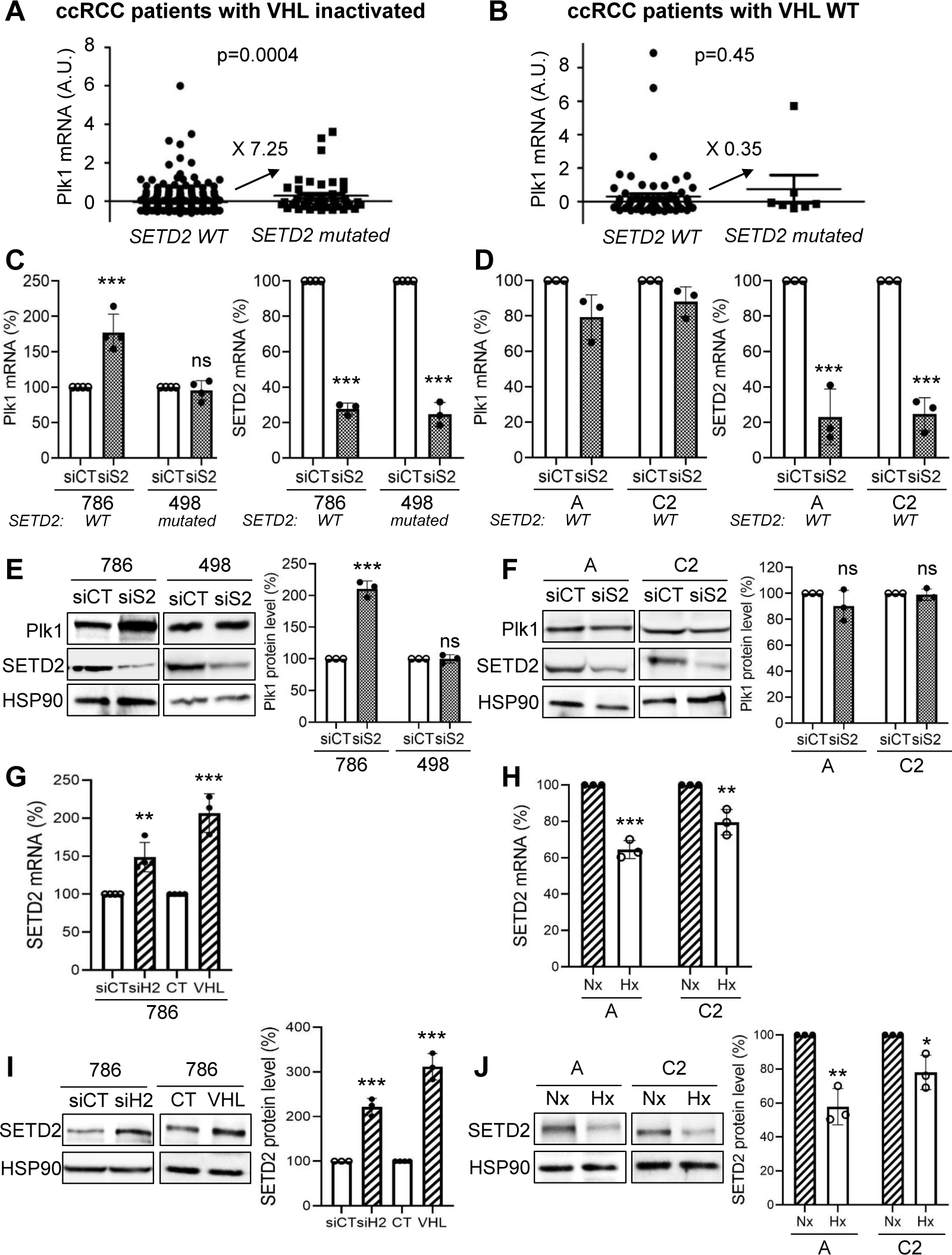
SETD2 inactivation induced Plk1 expression in ccRCC cells with inactivated VHL. **A and B**, Levels of Plk1 mRNA (z-score) in ccRCC patients with wild-type SETD2 were compared to the levels in ccRCC patients with inactivated SETD2, in RCC patients with inactivated VHL (**A**) or in ccRCC patients with wild-type VHL (**B**). **C to F**, VHL-inactivated 786 and 498 ccRCC cell lines (**C, E**), or A and C2 ccRCC cells wild-type VHL (**D, F**) were transfected with siRNA against SETD2 (S2) for 72 h. The Plk1 and SETD2 mRNA levels were determined by qPCR (**C, D**). Plk1 and SETD2 expression was evaluated by immunoblotting. HSP90 served as a loading control (**E, F**). **G and H**, 786 cells (VHL-inactivated) were transfected with H2 siRNA for 48 hours or an expression vector coding for VHL (stable expression, 786+VHL). SETD2 mRNA levels were determined by qPCR (**G**). SETD2 expression was evaluated by immunoblotting. HSP90 served as a loading control. The quantification of Plk1 expression (mean of three experiments) is shown (**H**). **I and J**, ccRCC cell lines (VHL-WT), A and C2 cells were cultured in normoxia (Nx) or hypoxia 1% O2 (Hx) for 24 h. The SETD2 mRNA levels were determined by qPCR (**I**). SETD2 expression was evaluated by immunoblotting. HSP90 served as a loading control. The quantification of Plk1 expression (mean of three experiments) is shown (**J**). The value of the control condition was considered as the reference value (100). Results are represented as means of three or more independent experiments (biological replication) ± SEM. Statistics were analyzed using an unpaired Student’s *t* test: * p<0.05, ** p<0.01, *** p<0.001.

### Plk1 promotes an invasive phenotype and induces sunitinib resistance

The link between Plk1 and ccRCC aggressiveness was assessed through the analysis of the TCGA data base. A volcano plot showed 933 up-regulated (4.3%) and 316 down-regulated (1.5%) genes in tumors expressing high or low levels of Plk1 (Supplementary Fig. S5A). Hierarchical cluster analyses showed distinguishable expression profiles for tumors expressing high and low levels of Plk1 (Supplementary Fig. S5B). Pathway analysis showed that high levels of Plk1 positively correlated with high proliferation, strong invasive potential and resistance to p53-dependent cell death (Supplementary Fig. S5C and D).

To confirm the role of Plk1 in the aggressiveness of ccRCC, we generated 786 cells over-expressing *Plk1* (786 *Plk1-1* and 786 *Plk1-2*) (Fig. 4A). Over-expression of *Plk1* enhanced cell migration (Fig. 4B). Sunitinib-treated naive ccRCC cells exhibited characteristics of senescence, inhibition of cell proliferation, G1-S cell cycle arrest and DNA damage response attributed to p53 activation (19). The viability and death of cells over-expressing Plk1 were affected to a lesser extent by sunitinib (Fig. 4C and D). Sunitinib activated p53 (total and phosphorylated form (p-p53)) in 786 but not in 786 *Plk1-1* and 786 *Plk1-2* cells (Fig. 4E). Plk1 expression was increased in tumor samples of patients treated by sunitinib in a neo-adjuvant setting (20) (Supplementary Fig. S6A) through sunitinib-dependent hypoxia and/or by selecting sunitinib-resistant cells.

**Figure 4.**
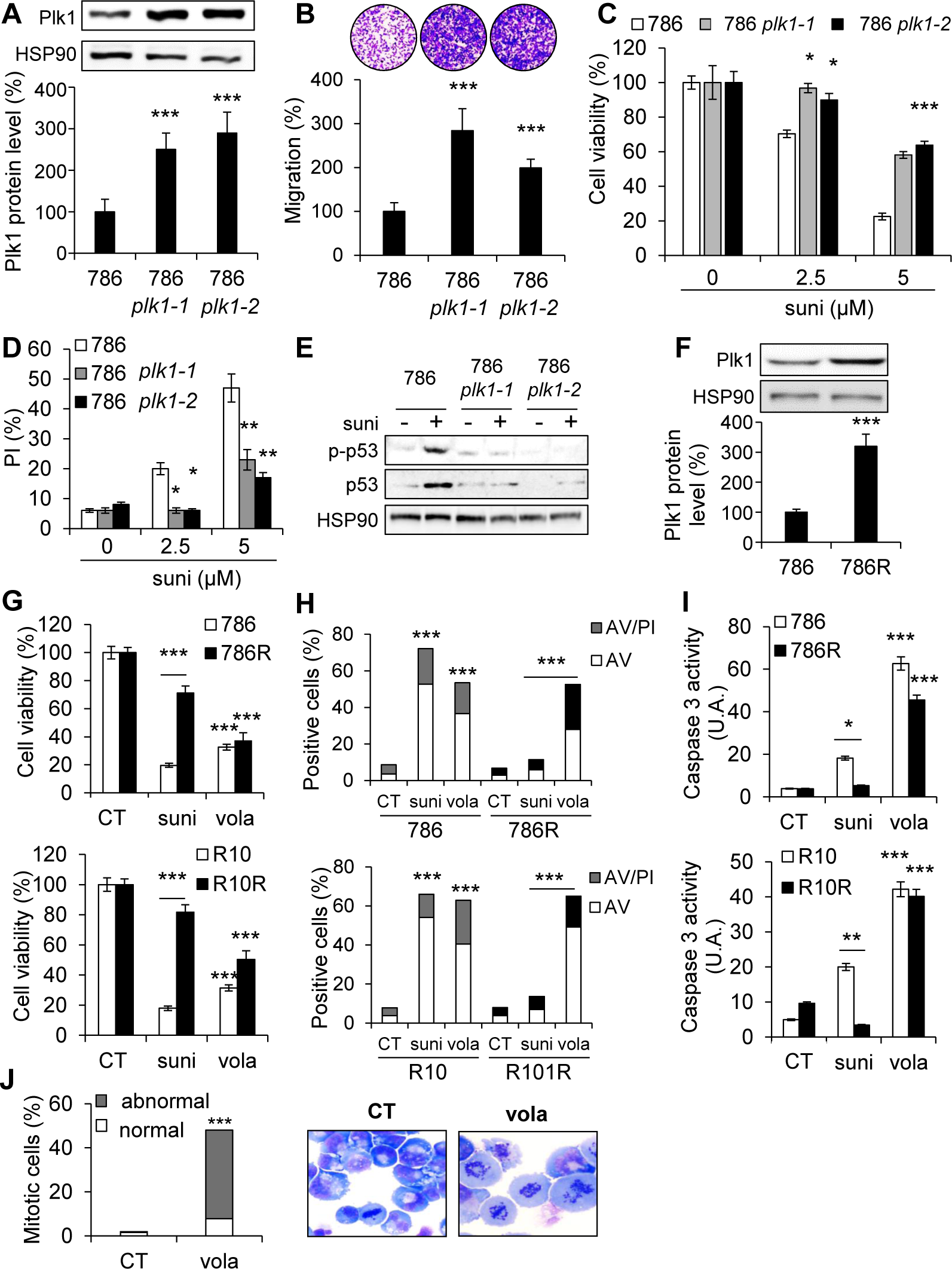
Plk1 over-expression induced aggressiveness, resistance to sunitinib and its inhibition by volasertib induced cell death. **A**, 786 cells were transfected with the *Plk1* expression vector and two clones (786 *Plk1-1* and 786 *Plk1-2*) stably expressing Plk1 were selected. Plk1 expression was evaluated by immunoblotting. HSP90 served as a loading control. The quantification of Plk1 expression (mean of three experiments) is shown. The value of the conditions with 786 cells were considered as the reference value (100). **B**, Serum-stimulated cell migration was analyzed using Boyden chamber assays on 786, 786 *Plk1-1* and 786 *Plk1-2* cells. The level of migration of 786 cells was considered as the reference value (100 %). Representative images of the lower surface of the membranes are shown. **C and D**, 786, 786 *Plk1-1* and *786 Plk1-2* cells were treated with 2.5 or 5 µM sunitinib (suni) for 48 h. Cell viability was measured with the XTT assay (**C**). Cell death was evaluated by flow cytometry. Cells were stained with PI. Histograms show PI-positive cells (**D**). **E**, 786, 786 *Plk1-1* and 786 *Plk1-2* cells were treated with 2.5 µM suni for 48 h. p-p53 and p53 expression were evaluated by immunoblotting. HSP90 served as a loading control. These results are representative of three independent experiments. **F**, Plk1 expression was evaluated by immunoblotting in 786 and 786 cells resistant to sunitinib (786R). HSP90 served as a loading control. The quantification of Plk1 expression (mean of three experiments ± SEM) is shown. Plk1 expression in 786 cells served as the reference value (100 %). **G to I**, 786 and 786R or R10 and R10R cells were treated with 100 nM volasertib (vola) or 5 µM sunitinib (suni) for 48 h. Cell viability was measured with XTT assays (**G**). Cell death was evaluated by flow cytometry. Cells were stained with PI and AV. Histograms show AV^+^/PI^−^ cells (apoptosis) and AV^+^/PI^+^ cells (post-apoptosis or another cell death) (**H**). Caspase-3 activity was evaluated using Ac-DEVD-AMC as a substrate (**I**). **J**, 786 cells were treated with 100 nM vola for 24 h. Hematoxilin and Eosin (HE) staining was assessed and the number of cells with normal and abnormal mitosis was evaluated. Results are represented as means of three or more independent experiments (biological replication) ± SEM. Statistics were determined using an unpaired Student’s *t* test: * p<0.05, *** p<0.001.

Plk1 expression and sunitinib resistance relationship was further addressed in sunitinib-resistant cells (786R). Plk1 expression was higher in 786R (Fig. 4F). p38 MAP Kinase (p38) activity is a key player driving resistance to sunitinib (20). Therefore, we hypothesised that p38 was involved in Plk1 expression. A p38 inhibitor decreased Plk1 mRNA and protein levels in 786R cells (Supplementary Fig. S6B and C). These results suggest that Plk1 is a key player in resistance to sunitinib by bypassing senescence and by promoting dissemination capabilities.

### Volasertib induced the death of resistant ccRCC cells and of primary ccRCC cells

Since Plk1 is key in ccRCC aggressiveness, we examined the sensitivity of ccRCC cells to different Plk1 inhibitors. ccRCC cell lines and primary ccRCC cells were more sensitive to Plk1 inhibitors than normal kidney cells (Supplementary Table S3). At low concentrations, volasertib decreased the proliferation and induced the death of ccRCC cells (Supplementary Fig. S6D and E). Volasertib-mediated cell death was mainly due to mitotic catastrophe (MC, Fig. S6*F*). Induction of MC was confirmed by hematoxilin-eosin (HES) staining and morphological analysis. Volasertib decreased cell viability (Fig. 4G), clonogenic potential (Supplementary Fig. S6D), and induced apoptosis through caspase 3 activation, increased abnormal mitosis and cytokinesis in sunitinib-sensitive and-resistant cells (Fig. 4H-J).

Volasertib had no effect on normal kidney cells but it decreased the viability, the clonogenic potential, induced MC leading to cell death and caspase 2 activation of primary ccRCC cells (Fig. 4K-M; Supplementary Fig. S7).

These results suggest that Plk1 inhibition bypasses resistance to sunitinib and are highly efficient in primary ccRCC cells.

### Volasertib has a strong anti-tumor effect in experimental ccRCC in mice, in a model of metastasis in the zebrafish and on primary tumor fragments

Volasertib inhibited the growth of experimental tumors in mice (Fig. 5A and B) more efficiently than sunitinib (Supplementary Fig. S8A). Control tumors (CT) were heavier than tumors from volasertib-treated mice, in which mitotic defects (HES, Supplementary Fig. S8B) and decreased numbers of proliferative cells (Ki67 staining) were observed (Fig. 5C). Volasertib decreased the number of blood vessels reaching the tumors, and their density (Fig. 5D and E; Supplementary Fig. S7C). These results suggest that volasertib is an angiogenesis inhibitor.

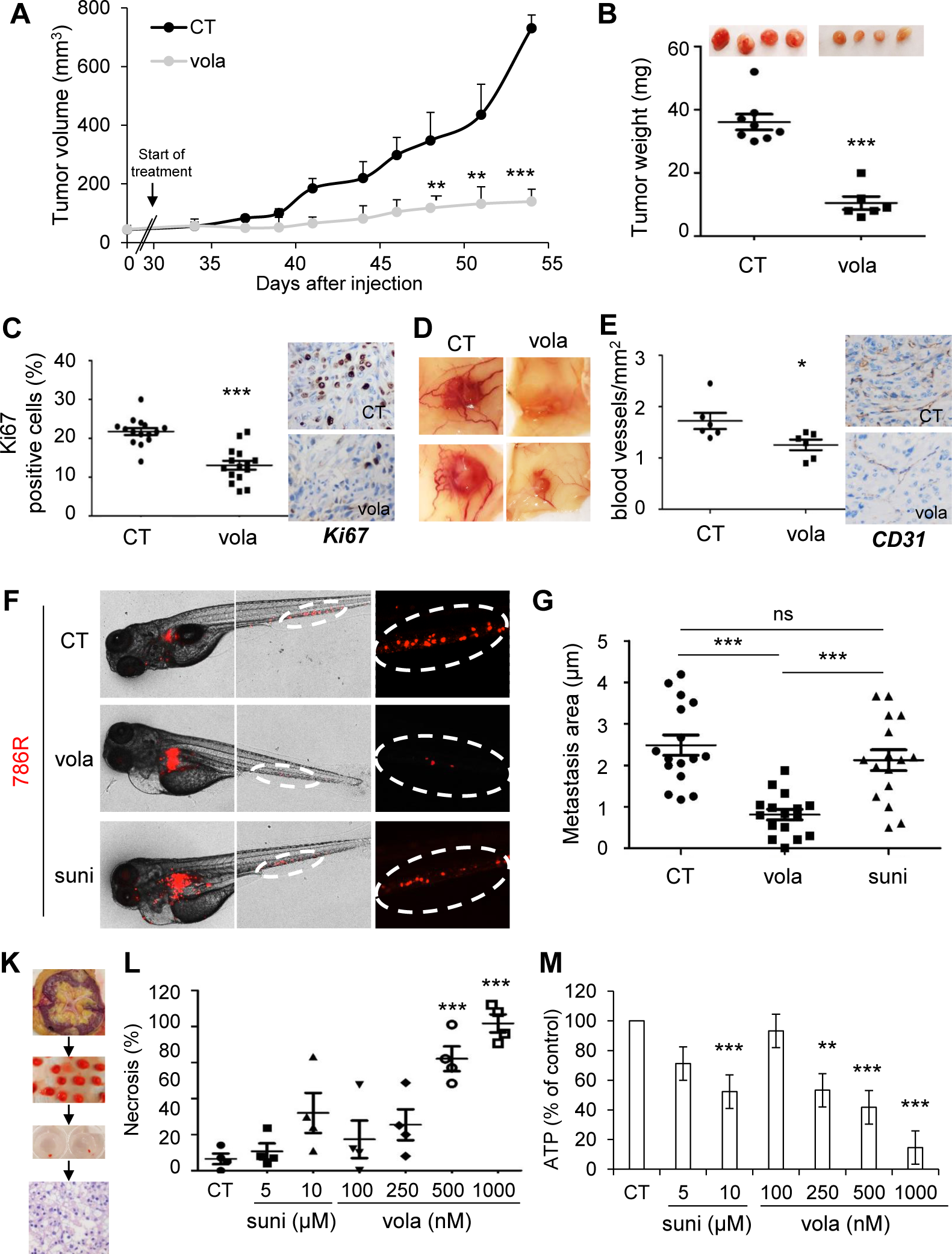
Volasertib inhibited the growth of experimental ccRCC in mice, decreased metastasis in zebrafish model and induced the death of 3D ccRCC primary tumors. **A to E**, 7.10^6^ 786 cells were subcutaneously injected into nude mice (*n*=8 per group). 30 days after injection, all mice developed tumors and were treated with the control solution or 25 mg/kg volasertib (vola) by gavage twice a week. **A**, The tumor volume was measured twice weekly as described in the materials and methods. **B**, The tumor weight at the end of the experiment. **C**, IHC of KI67 (proliferative cells). Representative images are shown. **D**, Representative images of tumors with blood vessels are shown. **E**, IHC of CD31 (blood vessels). Representative images are shown. **F and G**, Zebrafish embryos (n=45) were injected with 786R (labelled with red DiD) into the perivitelline space. 24 hours later, only zebrafish with metastasis are chosen and treated for 48h with sunitinib (suni, 1 μM) or vola (50 nM). Zebrafish embryos were monitored for investigating tumor metastasis using a fluorescent microscope. Representatives images are shown (**F**) and area of metastasis are quantified (**G**). **K to M**, A sample of tumors following nephrectomy of the patient was analyzed by a pathologist (4 ccRCC patients). The tumor sample was then cut into fragments of about 5mm^3^, cultured in a specific medium and treated for 72 hours with sunitinib (suni) or vola (**K**). Tumor fragments were paraffin-embedded and stained with HES to quantify the areas of necrosis (**L**). Tumor fragments were lyzed, and the concentration of ATP determined to provide a read-out of the tumor fragment viability (**M**). Statistics were determined using an unpaired Student’s *t* test (A, B, C, E) or Annova analysis (Bonferroni’s comparison, G, L, M): * p<0.05, ** p<0.01, *** p<0.001

Zebrafish were used as an elegant and pertinent model of metastasis by assessing dissemination of tumor cells from the site of injection to the tail (21). In this model, 786R had a strong ability to metastasize. While sunitinib was unable to inhibit distant metastases (in the tails) of the zebrafishes, vola reduced significantly their size and number (Fig. 5F and G).

For a theranostic approach, volasertib efficacy was tested on sections of tumors obtained from surgical specimens (Fig. 5K). HES staining showed necrosis after treatments (Fig. 5L; Supplementary Fig. S8D). Sunitinib and volasertib decreased the viability of tumor fragments but only volasertib induced necrosis (Fig. 5M and L). These results support the relevance of volasertib as a therapeutic alternative for ccRCC.

### Plk1 expression and molecular ccRCC subtypes as indicator for therapy decision

Analysis of the TCGA database and the Cancer Immunome Atlas (TCIA)) showed that M1 ccRCC patients with a low expression of Plk1 and PDL1 had the longest OS, patients with a high expression level of PDL1 had an intermediate OS, and patients with high Plk1 and low PDL1 had the shortest OS (Supplementary Fig. S9A). PDL1 expression has been associated with good response to immunotherapy in patients with metastatic ccRCC (22). The Immunoscore determined by the TCIA support these clinical observations. Plk1 over-expression and the low expression of PDL1 were associated with a bad immunoscore reflecting a poor response to immunotherapy (Supplementary Fig. S9B). The impact of Plk1 expression on PFS in TKI first-line treatment (sunitinib, pazopanib and sorafenib, Table 3A) was investigated on 58 primary ccRCCs. Plk1 expression was increased in tumors of patients with an intermediate and poor IMDC (International Metastatic RCC Database Consortium) score (Fig. 6A). Patients with an intermediate and poor IMDC score are poorer responders to TKI. According to *in vitro* results, Plk1 over-expression induced resistance to TKI (PFS of 3.5 months *vs* 14 months, p=0.0004, Fig. 6B) and particularly to sunitinib (PFS of 7 months *vs* 20 months, p=0.0157, Fig. 6C). Plk1 levels subclassified two categories of patients with an intermediate IMDC score. Low expression of Plk1 was associated with a better outcome on TKI in this heterogeneous group (PFS of 16 months *vs* 3 months, p=0.0137, Fig. 6D). Plk1 over-expression is associated with a shorter OS following TKI (OS of 12 months *vs* 34 months, p=0.022, Fig. 6E). Multivariate analyses showed that Plk1 mRNA levels are indicative of PFS for patients on TKI independently of the IMDC score (Table 1). While a bad IMDC score was indicative of PFS (Table 1B), it lost its significance in a multivariate analysis including Plk1 (Table 1C). These results suggest that Plk1 expression orient the therapeutic decision in addition to clinical parameters. Plk1 expression was increased in ccrcc1&4 in comparison to ccrcc2&3 molecular subtypes (Fig. 6F). SETD2 mutations are mostly present in the ccrcc1 subtype (Figure S7 in (10)). Thus, SETD2 mutations and Plk1 expression seem to be frequent in ccrcc1-subtype tumors. Hence, patients of this subtype, which responds poorly to TKIs and immunotherapy, are good candidates for Plk1 targeting agents such as volasertib.

**Table 1.** The characteristics of the metastatic ccRCC patients included in the study and a multivariate analysis. **A**, Patient characteristics and univariate analysis with the Fisher or Ki^2^ test. Statistical significance (*p* values) is indicated. **B**, Multivariate analysis of the IMDC score and PFS or OS. The multivariate analysis was performed using Cox regression adjusted to the IMDC score. Statistical significance (*p* values) is indicated. **C**, Multivariate analysis of Plk1, the IMDC score and PFS. The multivariate analysis was performed using Cox regression adjusted to the IMDC score. Statistical significance (*p* values) is indicated. (see Fig.6).

|  | Total | Low Plk1 | High Plk1 | p value |
| --- | --- | --- | --- | --- |
| **Number** | 58 | 43 | 15 |  |
| **pM** |  |  |  |  |
| 1 | 58 (100%) | 43 (100%) | 15 (100%) |  |
| **Fuhrman grade** |  |  |  |  |
| 2 | 1 (1.7%) | 0 (0%) | 1 (6.7%) | 0.0005 |
| 3 | 23 (39.7%) | 23 (53.5%) | 0 (0%) |  |
| 4 | 34 (58.6%) | 20 (46.5%) | 14 (93.3%) |  |
| **Heng score (IMDC score)** |  |  |  |  |
| good | 7 (12%) | 7 (16.3%) | 0 (0%) | 0.234 |
| intermediate | 39 (67.2%) | 28 (65.1%) | 11 (73.3%) |  |
| bad | 12 (20.8%) | 8 (18.6%) | 4 (26.7%) |  |
| **Molecular subtype** |  |  |  |  |
| ccrcc1 | 16 (27.6%) | 9 (20.9%) | 7 (46.7%) | 0.0005 |
| ccrcc2 | 30 (51.7%) | 29 (67.5%) | 1 (6.7%) |  |
| ccrcc3 | 3 (5.2%) | 1 (2.3%) | 2 (13.3%) |  |
| ccrcc4 | 9 (15.5%) | 4 (9.3%) | 5 (33.3%) |  |
| **1<sup>st</sup> line treatment** |  |  |  |  |
| sunitinib | 39 (67.2%) | 29 (67.4%) | 10 (66.7%) | 0.277 |
| pazopanib | 13 (22.4%) | 11 (25.6%) | 2 (13.3%) |  |
| sorafenib | 6 (10.3%) | 3 (7%) | 3 (20%) |  |
| **PFS (months) / progression %** | 11<br>84% | 14<br>79% | 4<br>100% | 0.004 |
| **OS (months) / Death %** | 21<br>79% | 34<br>77% | 12<br>93% | 0.022 |

B
| IMDC score | Description | Mediane (months) | coef | HR | p value |
| --- | --- | --- | --- | --- | --- |
| PFS | good | 81 |  |  |  |
|  | Intermediate | 14 | 1.264 | 1.874 | 0.240 |
|  | poor | 3 | 2.078 | 3.054 | 0.037 |
| OS | good | NR |  |  |  |
|  | Intermediate | 22 | 1.279 | 3.538 | 0.083 |
|  | poor | 6.5 | 2.063 | 7.292 | 0.007 |

C
| PFS 1<sup>st</sup> line TKI | Description | coef | HR | p value |
| --- | --- | --- | --- | --- |
| Biological parameter<br>**Plk1** |  | 0.334 | 1.397 | 0.008 |
| Clinical parameters<br>**IMDC score** | good<br>Intermediate<br>poor | 0.171<br>0.909 | 1.187<br>2.482 | 0.764<br>0.141 |

**Figure 6.**
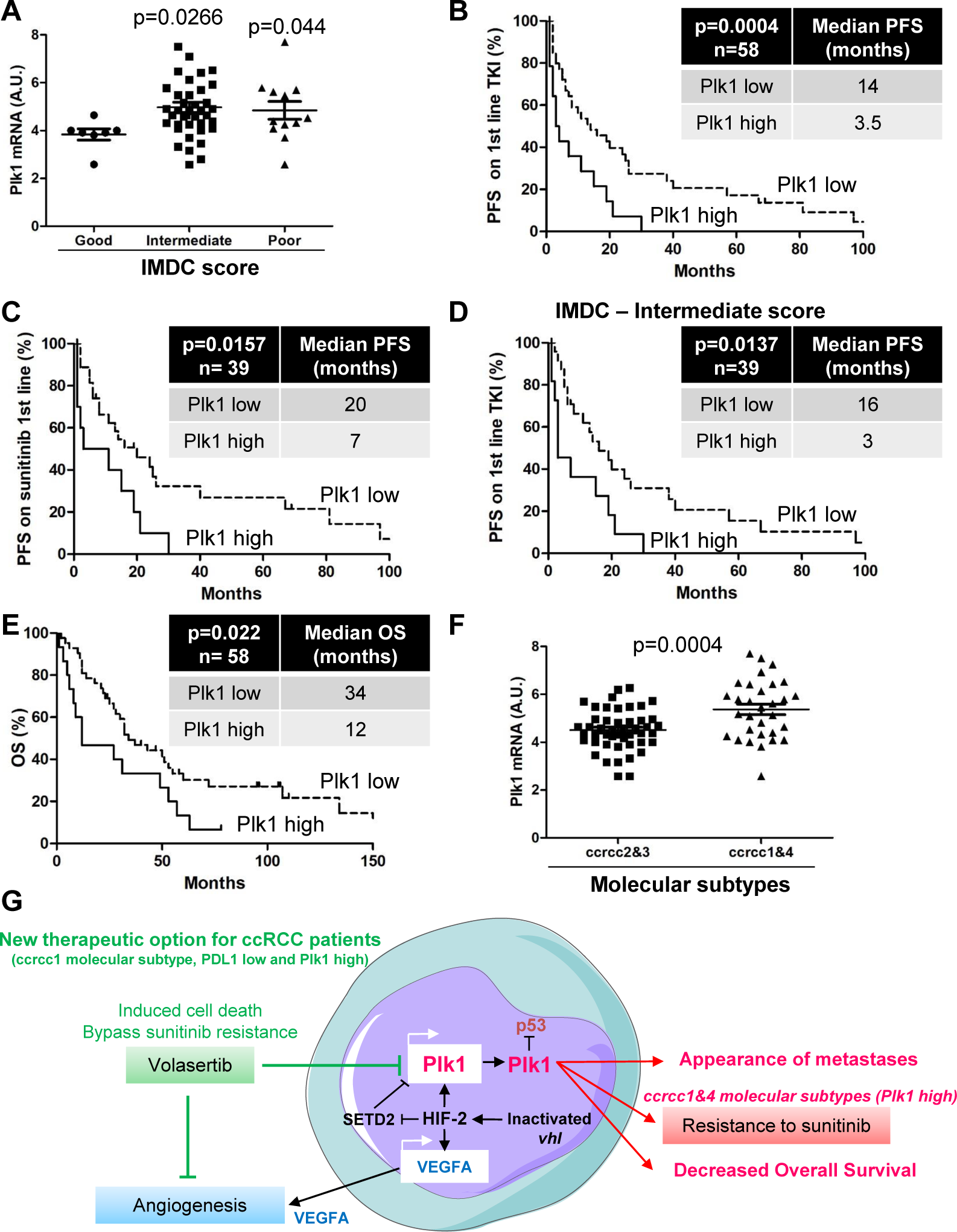
Plk1 is associated with resistance to fist-line VEGFR-TKI treatment in ccRCC. The tumors of 58 metastatic ccRCC patients were analyzed for the Plk1 mRNA level. **A**, The levels of Plk1 mRNA in tumors from patients with a good, intermediate and poor IMDC score were compared. **B to E**, The levels of Plk1 mRNA in tumors of 58 metastatic ccRCC patients treated with VEGFR-TKI in the first line correlated with PFS (**B**) or with OS (**E**). The levels of Plk1 mRNA in tumors of 27 metastatic ccRCC patients treated with sunitinib in the first line correlated with PFS (**C**). The levels of Plk1 mRNA in tumors from patients with an intermediate IMDC score correlated with PFS (**D**). (**F**) The levels of Plk1 mRNA in tumors of the ccrcc2&3 and ccrcc1&4 subtypes were compared. The third quartile of Plk1 expression was chosen as the cut-off value. Statistics were determined using an unpaired Student’s *t* test. The Kaplan-Meier method was used to produce survival curves and analyes of censored data were performed using Cox models. Statistical significance (p values) is indicated. (see Table 1).

## Discussion

Plk1 is known to be upregulated in highly proliferative tumors. However, its regulation in cancer cells are poorly understood. Highly proliferative tumors often experience a high-grade of hypoxia owing to the rapid expansion of the tumor mass. This would create a discordant scenario between severe hypoxia and high proliferation of tumor cells. How could tumor cells continue to proliferate at a high rate in the presence of severe hypoxia? At this time of writing, there are no mechanisms to satisfactorily explain this interesting phenomenon. However, several hypoxia-related characteristics are associated with fast tumor growth, including: 1) Fast-growing tumors often encounter necrosis owing to deprivation of oxygen; 2) Hypervascularization in tumors by a hypoxia-induced VEGF mechanism; 3) Ineffective drug responses due to poor delivery of cancer drugs; 4) Highly metastatic because of hypoxia-driven cancer stem cell seeding mechanism; 5) High expression of growth factors and cytokines through a hypoxia-regulated mechanism; 6) Alteration of TME; and 7) Reprogramming of cancer cell metabolism. Conversely, these features may serve as a predictive marker to dictate tumor hypoxia. In this study, we provide an example of a hypoxia-regulated oncogenic protein that significantly contributes to cancer metastasis and drug responses.

Most hypoxia-regulated genes are mediated by HIF-1α, which is a master regulator of hypoxia-triggered responses. For example, hypoxia-induced VEGF and EPO expression is controlled by HIF-1α (23). In additional to environmental hypoxia, genetic mutations can also trigger a similar hypoxia-like response in cancer cells. In this report, we focus our study on ccRCC that often carries a mutated and dysfunctional VHL. In the absence of VHL, HIF-α increased target genes irrespective of the oxygen concentration. Although HIF-1α and HIF-2α bind to similar sequences, they play independent roles (30). HIF-1 drives genes involved in metabolism, whereas HIF-2 stimulates genes coding for pro-survival factors. Therefore, HIF-1α is tumor suppressor whereas HIF-2α is an oncogene in ccRCC (31,32). We demonstrated that hypoxia-dependent up-regulation of Plk1 depends on a HIF-2-dependent stimulation of transcription, and mutation of SETD2 enhanced it. Surprisingly, Plk1 is a target gene for HIF-2a, but HIF-1a in human ccRCC, suggesting the existence of an alternative mechanism in driving tumor growth and metastasis. Although not investigated in this study, activation of HIF-2a may induce expression of a variety of growth factors and cytokines, which collaboratively promote oncogenesis with Plk1. At the advanced stage of tumor development, genetic mutation-triggered hypoxia-like response and environmental hypoxia play dual role in driving cancer progression. We provide clinical evidence to supportive this hypothesis by showing high Plk1 expression in necrotic, larger, and poor prognosis tumor. These findings show that Plk1 is a central player for facilitating tumor development and progression. Metastatic ccRCC patients relapse despite the introduction of angiogenesis inhibitors (VEGFR-TKI) and immune therapy. Predictive markers relevant to current treatments and new therapeutic targets are required. In addition to VEGFR, sunitinib directly targets tumor cells to inhibit cell proliferation, migration, and survival. As malignant cells are genetically unstable, ccRCC patients receiving with sunitinib treatment often develops resistance through a compensatory mechanism of survival. In supporting this line of thinking, sunitinib-resistant ccRCC exhibit higher Plk1 expression, suggesting that Plk1 may play a crucial role in developing sunitinib resistance. If so, blocking Plk1 provides an attractive and alternative therapeutic regimen for treating sunitinib resistant ccRCC. Indeed, we provide compelling evidence to show that sunitinib resistant ccRCC are highly sensitive to Plk1 inhibition. This exciting finding warrants clinical validation.

We provided evidence that Plk1 is involved in resistance to sunitinib by bypassing p53-dependent senescence (19). An imbalance in the interactions between these two proteins and the resulting deregulation of oncogenic pathways contributes to cancer development. Epithelial to mesenchymal transition (EMT) enables cancer cells to avoid apoptosis, anoïkis, and oncogene addiction (24). Over-expression of Plk1 increases cell migration, a key process induced during EMT. Our results are consistent with the down-regulation of epithelial markers and up-regulation of mesenchymal markers in prostate epithelial cells over-expressing Plk1 (25).

Plk1 is associated with resistance to doxorubicin, paclitaxel, and gemcitabine (26). Targeting the addiction to Plk1 appears relevant to increase the sensitivity to chemotherapy (13, 14), which is consistent with volasertib-dependent ccRCC cell death in sunitinib sensitive and -resistant cells. Plk1 was described as a therapeutic target for ccRCC by an independent approach and a non-clinically approved Plk1 inhibitor (BI 2536) inhibited the growth of experimental tumors (27). We further explored the molecular mechanism linking over-expression of Plk1 and ccRCC aggressiveness. The link between two major cancer hallmarks, cell proliferation through activation of Plk1 and hypoxia through HIF-α stabilisation constitutes the main breakthrough of the present study. ccRCC represents a paradigm for HIF-dependent tumor aggressiveness. The generalisation of this concept to different tumors (breast, liver, lung, pancreatic cancers, melanoma and sarcoma) brings added value to improve the treatment of these cancers.

Targeting Plk1 inhibited tumor and endothelial cell proliferation in mice model and development of metastasis in zebrafish model. For the first time, we show that ccRCC cells can metastasize in zebrafish without genetic modification beforehand. Therefore, the therapeutic efficacy of Plk1 inhibitors also rely on the inhibition of angiogenesis, a key phenomenon in ccRCC.

Plk1 is a driver of tumor growth orchestrated by the HIF-2 oncogenic pathway. Our study linked Plk1 to shorter survival in both M0 and M1 patients. Plk1 is a prognostic factor independent of Fuhrman grade. Hence, a biological marker independent of clinical parameters provides added value to the management of patients.

The gold-standards for metastatic ccRCC patients in the first-line are TKI, immunotherapy (anti-PD1 + anti-CTLA4, (28)) or a combination of both therapies (29, 30).

The clinical parameters of the IMDC score are poorly informative for patients in the intermediate group. The four subtypes ccrcc1–4, based on biological parameters refine the therapeutic strategy (10). ccrcc1-tumors have an immune-cold indicative of immunotherapy refractoriness and a bad response to TKI. Tumors of the ccrcc1 subtype strongly express Plk1. High Plk1 mRNA levels correlated with a poor response to immunotherapy (31). Therefore, Plk1 inhibitors represent a relevant strategy for these patients. The following nomogram appears decisional for the therapeutic strategy for patients of the different subgroups:

- ccrcc2&3 subtypes (low PDL1 and Plk1 expression) are eligible for TKI,
- ccrcc4 subtype (high PDL1 and Plk1 expression) are eligible for immunotherapy,
- ccrcc1 subtype (low PDL1 expression but strong Plk1 expression) are eligible for treatment with Plk1 inhibitors

Our study deciphered the phenomenon linking a physical driver of tumor aggressiveness (hypoxia) to a biological determinant of tumor cell proliferation and angiogenesis (Plk1). The link between the two actors is HIF-2, which drives *Plk1* gene transcription through SETD2-dependent chromatin-remodelling. In addition to its tumor promoting role, Plk1 drives resistance to TKI and appears as a key target for a subgroup of metastatic ccRCC patients in therapeutic impasses (Fig. 6G).

## Materials and methods

### Reagents and antibodies

Inhibitors were purchased from Selleckchem. Anti-HSP90 and anti-tubulin antibodies were purchased from Santa Cruz Biotechnology. Anti-Plk1 antibodies were purchased from Abcam. The anti-HIF-2α antibody was purchased from Novus Biochemicals. The rabbit polyclonal anti-HIF-1α antibodies were previously described (32).

### Cell culture

RCC4 (R4), ACHN (A), Caki-2 (C2), 786-0 (786) and A498 (498) ccRCC cell lines were purchased from the American Tissue Culture Collection. RCC10 (R10) was a kind gift from Dr. W.H. Kaelin (Dana-Farber Cancer Institute, Boston, MA). Primary cells were already described and cultured in a medium specific for renal cells (PromoCell, Heidelberg Germany) (18). 786R and RCC10R were previously described (33). An INVIVO2 200 anaerobic workstation (Ruskinn Technology Biotrace International Plc) set at 1 % oxygen, 94 % nitrogen, and 5 % carbon dioxide was used for hypoxic conditions (34).

### Patients

Informed consent was obtained from all individual participants included in the study. All patients gave written consent for the use of tumor samples for research. The study included only the major patients. This study was conducted in accordance with the Declaration of Helsinki.

Primary tumor samples of M0 ccRCC patients were obtained from the Rennes (35) and Bordeaux University Hospitals and UroCCR group (Fig. 1 and Supplementary Table 2A). Paraffin embedded Samples of primary tumors from M1 ccRCC patients were obtained from Leuven Hospital (Fig. 6 and Table 1). Plk1 mRNA levels of were measured using a customized Nanostring Counter(c) gene panel. The DFS, PFS and OS were calculated from patient subgroups with Plk1 mRNA levels that were less or greater than the third quartile value.

### siRNA assay

siRNA transfection was performed using Lipofectamine RNAiMAX (Invitrogen). Cells were transfected with either 50 nM of siHIF-1α (siH1) (36) and/or HIF-2α (siH2) (37) or sicontrol (siCT, Ambion, 4390843). Two days later, experiments were performed as described above.

### Quantitative Real-Time PCR (qPCR) experiments

1 µg of total RNA was used for the reverse transcription, using the QuantiTect Reverse Transcription kit (QIAGEN), with blend of oligo (dT) and random primers to prime first-strand synthesis. SYBR master mix plus (Eurogentec) was used for qPCR. The mRNA level was normalized to 36B4 mRNA.

### Luciferase assays

Transient transfections were performed using 2 µl of lipofectamine (GIBCO BRL) and 0.5 µg of total plasmid DNA-renilla luciferase in a 500 µl final volume. The firefly control plasmid was co-transfected with the test plasmids to control for the transfection efficiency. 24 hours after transfection, cell lysates were tested for renilla and firefly luciferase. All transfections were repeated four times using different plasmid preparations. LightSwitch™ Promoter Reporter Plk1 was purchased from Active motif.

### Chromatin immunoprecipitation (ChIP)

These experiments were performed as already described (38). Briefly, cells were grown in normoxia or hypoxia (1% O2) for 24 hours (5–10 × 10^6^ cells were used per condition). Cells were then fixed with 1% (v/v) formaldehyde (final concentration) for 10 min at 37°C and the action of the formaldehyde then stopped by the addition of 125 mM glycine (final concentration). Next, cells were washed in cold PBS containing a protease inhibitor cocktail (Roche), scrapped into the same buffer and centrifuged. The pellets were re-suspended in lysis buffer, incubated on ice for 10 min, and sonicated to shear the DNA into fragments of between 200 and 1,000 base pairs. Insoluble material was removed by centrifugation and the supernatant was diluted 10-fold by addition of ChIP dilution buffer and pre-cleared by addition of salmon sperm DNA/protein A agarose 50% slurry during 1 hours at 4°C. About 5% of the diluted samples was stored and constituted the input material. Immunoprecipitation was then performed by addition of anti-HIF-2α or anti-tubulin as IgG control antibodies for 24 hours at 4 °C. Immuno-complexes were recovered by adding 50 % of salmon sperm DNA/protein A agarose and washed sequentially with low salt buffer, high salt buffer, LiCl buffer and TE. DNA complexes were extracted in elution buffer, and cross-linking was reversed by incubating overnight at 65 °C in the presence of 200 mM NaCl (final concentration). Proteins were removed by incubating for 2 hours at 42 °C with proteinase K and the DNA was extracted with phenol/chloroform and precipitated with ethanol. Immunoprecipitated DNA was amplified by PCR with the following primers:

Plk1 primers: Forward: 5’-AGTGAACCGCAGGAGCTTTC-3’, Reverse: 5’-TTAAAATCCAAACCCGCCCG-3’;

Positive control (PDH3) primers: Forward: 5’-TTCTCTGGTGACTGGGGTAGAGAT-3’, Reverse: 5’-GAGCCCATGCAATTAGGCACAGTA-3’;

Negative control (Ang2 – 9351) primers: Forward: 5’-TCACCTGAGGATACAGAGAC-3’, Reverse: 5’-AGCGACAGGCAAATCTATCCA-3’.

### Cell viability (XTT)

Cells (5×10^3^ cells/100 μl) were incubated in a 96-well plate with different effectors for the times indicated in the figure legends. 50 μl of sodium 3′-[1-phenylaminocarbonyl)-3,4-tetrazolium]- bis(4-methoxy-6-nitro) benzene sulfonic acid hydrate (XTT) reagent were added to each well. The assay is based on the cleavage of the yellow tetrazolium salt XTT to form an orange formazan dye by metabolically active cells. Absorbance of the formazan product, reflecting cell viability, was measured at 490 nm. Each assay was performed in quadruplicate.

### Cytospin preparations and Hematoxylin–Eosin staining

Cytospin preparations were also obtained using the cytocentrifuge (Thermo Scientific Cytospin 4, Thermo, Pittsburgh, PA, U.S.A.) at 900 rpm for 9 min. Smears were stained with hematoxylin– eosin (HES), for morphological assessment.

### Measurement of the caspase activity

After stimulation, cells were lyzed for 30 min at 4°C in lysis buffer (39), and lysates were cleared at 10,000 *g* for 15 min at 4°C. Each assay was done with 25 μg of protein. Cellular extracts were incubated in a 96-well plate with Ac-DEVD-AMC (caspase 3) or Ac-VDVAD-AMC (caspase 2) for various times at 37°C. Caspase activity was measured at 410 nm in the presence or absence of 1 μM of Ac-DEVD-CHO.

### Flow cytometry

#### Analysis of apoptosis

After stimulation, cells were washed with ice-cold PBS and stained with the annexin-V-fluos staining kit (Roche) according to the manufacturer’s procedure. Fluorescence was measured using the FL2 and FL3 channels of a fluorescence-activated cell sorter apparatus (FACS-Calibur cytometer).

#### Cell cycle analysis

After treatment, cells were washed, fixed in ethanol 70% and, finally, left overnight at −20°C. Cells were next incubated in PBS, 3 μg/ml RNase A and 40 μg/ml of propidium iodide (PI) for 30 min at 4 °C. Cellular distribution across the different phases of the cell cycle or DNA content was analyzed with a FACS-Calibur cytometer.

### Tumor xenograft experiments

*Ectopic model of ccRCC:* 7. 10^6^ 786 cells were injected subcutaneously into the flank of 5-week-old nude (nu/nu) female mice (Janvier, France). The tumor volume was determined with a caliper (v = L*l^2^*0.5). When the tumor reached 50 mm^3^, mice were treated five times a week for 4 weeks, by gavage with placebo (dextrose water vehicle), sunitinib (40 mg/kg) or twice a week for 4 weeks with volasertib (25 mg/kg). This study was carried out in strict accordance with the recommendations of the Guide for the Care and Use of Laboratory Animals. Our experiments were approved by the ‘‘Comité national institutionnel d’éthique pour l’animal de laboratoire (CIEPAL)’’ (reference: NCE/2015-255).

### Immunohistochemistry

Sections from blocks of formol-fixed and paraffin-embedded tumors were examined for immunostaining. Sections were incubated with monoclonal anti-mouse CD31 (clone MEC 13.3, BD Pharmingen, diluted at 1:500) or Ki67 (clone MIB1, DAKO, Ready to use) antibodies. Biotinylated secondary antibody (DAKO) was applied and binding was detected with the substrate diaminobenzidine against a hematoxylin counterstain.

### Zebrafish metastatic tumor model

All animal experiments were approved by the Northern Stockholm Experimental Animal Ethical Committee. Zebrafish embryos were raised at 28°C under standard experimental conditions. Zebrafish embryos at the age of 24 hpf were incubated in aquarium water containing 0.2 mmol/L 1-phenyl-2-thio-urea (PTU, Sigma). At 48-hpf, zebrafish embryos were dechorionated with a pair of sharp-tip forceps and anesthetized with 0.04 mg/mL of tricaine (MS-222, Sigma). Anesthetized embryos were subjected for microinjection. 786R tumor cells were labeled in vitro with a Vybrant DiD cell-labeling solution (LifeTechnologies). Tumor cells were resuspended in PBS and approximatively 5 nL of the cell solution were injected into the perivitelline space (PVS) of each embryo by an Eppendorf microinjector (FemtoJet 5247). Non-filamentous borosilicate glass capillaries needles were used for injection and the injected zebrafish embryos were immediately transferred into PTU aquarium water with treatment. 24 hours later, only zebrafish with metastasis were chosen and treated. Zebrafish embryos were monitored 48 hours for investigating tumor metastasis using a fluorescent microscope (Nikon Eclipse 90).

### Treatment of primary ccRCC tumors

A sample of the tumor obtained just after nephrectomy was provided by a pathologist. The tumor sample was then cut into pieces of about 5mm^3^ and cultured in a specific medium (18) and treated for 72 hours with sunitinib or volasertib. Tumor fragments were then paraffin-embedded and analyzed using HES and necrosis area were quantified. Tumor fragments were also lyzed, and the concentration of ATP represented a read-out of the viability of the tumor fragments.

### Statistical analysis

#### For in vitro and in vivo analysis

All data are expressed as the mean ± the standard error (SEM). Statistical significance and p values were determined by the two-tailed Student’s *t*-test. One-way ANOVA was used for statistical comparisons. Data were analyzed with Prism 5.0b (GraphPad Software) by one-way ANOVA with Bonferroni post hoc.

#### For patients

The Student’s *t*-test was used to compare continuous variables and chi-square test, or Fisher’s exact test (when the conditions for use of the *χ*^2^-test were not fulfilled), were used for categorical variables. To guarantee the independence of Plk1 as a prognostic factor, the multivariate analysis was performed using a Cox regression adjusted to the Fuhrman grade. DFS was defined as the time from surgery to the appearance of metastasis. PFS was defined as the time between surgery and progression, or death from any cause, censoring live patients and progression free at the last follow-up. OS was defined as the time between surgery and the date of death from any cause, censoring those alive at the last follow-up. The Kaplan-Meier method was used to produce survival curves and analyses of censored data were performed using Cox models. All analyses were performed using R software, version 3.2.2 (Vienna, Austria, https://www.r-project.org/).

## Data Availability

The data are included in the manuscript, details can be requested by email to the corresponding autor

## Author contributions

Investigation, MD, AV, LSC, PDN, XH, NN, WS, AH, ST, JP, AB, DA, JP; Methodology, MD; Resources; AV, NMM, BM, NRL, KB, AR, PA, JZR, SG, DB, BB, YC, JCB, DA; Conceptualization, MD, DA and GP; Statistical analysis, JV, RS, EC; Writing-original draft, YC, MD and GP; Supervision, Project administration and funding acquisition, MD and GP.

## Acknowledgements

This work was supported by the French Association for Cancer Research (ARC), the Fondation de France, the Ligue Nationale contre le Cancer (Equipe Labellisée 2019) the French National Institute for Cancer Research (INCA) and the FX Mora Foundation. This study was conducted as part of the Centre Scientifique de Monaco Research Program, funded by the Government of the Principality of Monaco. The samples from Bordeaux and associated data were collected, selected and made available within the framework of the project clinicobiological National Cancer Database Kidney UroCCR supported by l’Institut National du Cancer (INCa). We thank the IRCAN core facilities (animal and cytometry facilities) for technical help. We thank also the Department of Pathology, especially Arnaud Borderie and Sandrine Destree, for technical help.

## Supplementary Figure Legends

**Supplementary Figure S1.**
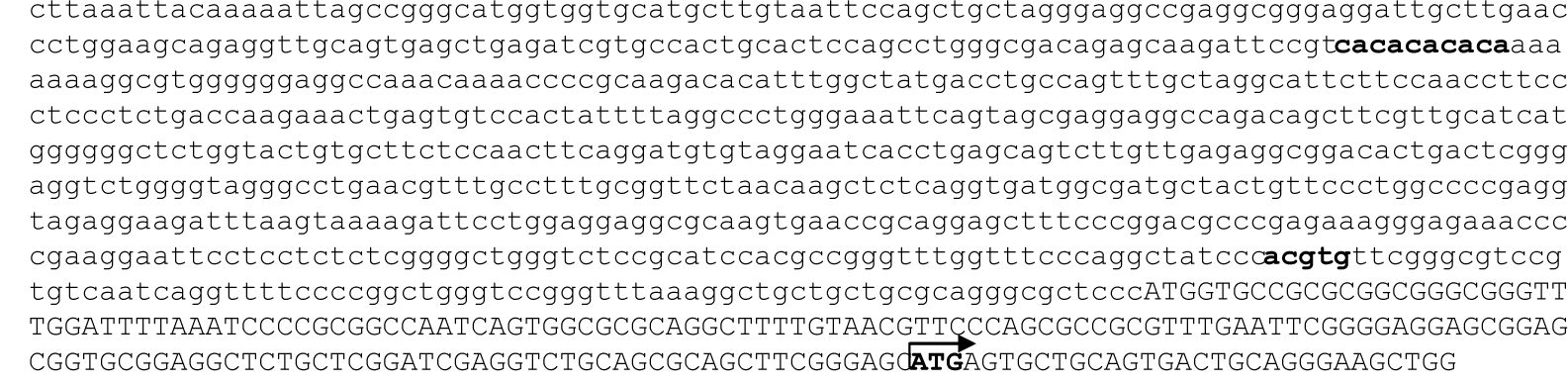
Presence of a HRE in the *Plk1* promoter determined by *in silico* analysis. HRE consensus sequence marked in bold.

**Supplementary Figure S2.**
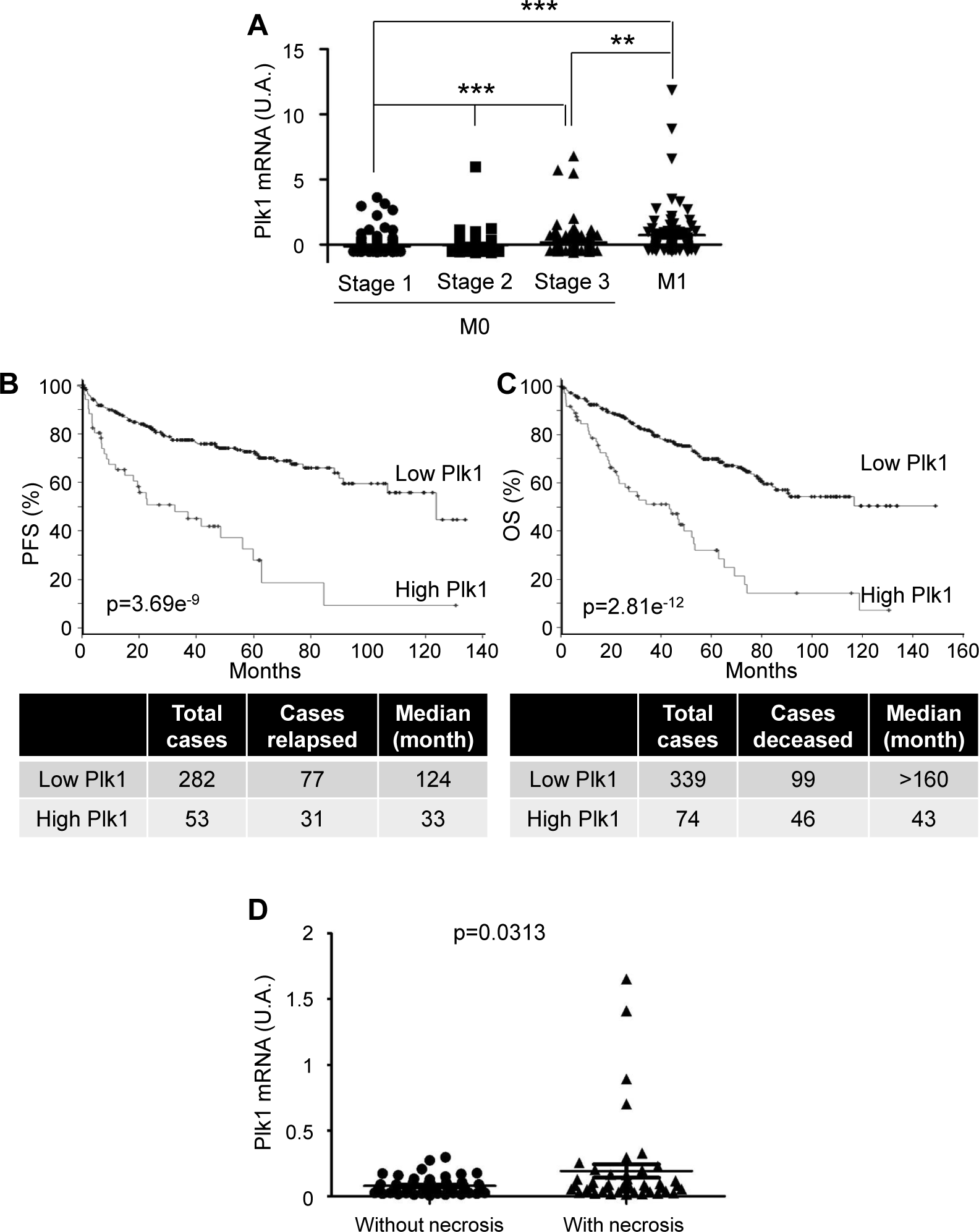
Plk1 is associated with poor prognosis in ccRCC. **A to C**, The tumors of ccRCC patients were analyzed for Plk1 mRNA levels. These results are in whole or in part based upon data generated by the TCGA Research Network. **A**, Analysis of the cBioportal database highlighted the levels of Plk1 mRNA in non-metastatic ccRCC (M0) stage 1, 2 or 3 or metastatic ccRCC (M1) patients. One-way ANOVA was used for statistical comparisons: ** p<0.01, *** p<0.001. **B and C**, The levels of Plk1 mRNA in tumors of ccRCC patients correlated with PFS (**B**) or with OS (**C**). PFS and OS were calculated from patient subgroups with mRNA levels that were less or greater than the median value. Statistical significance (p value) is indicated. **D**, 95 ccRCC patients were analyzed for Plk1 mRNA levels. The levels of Plk1 mRNA were compared in tumors with or without necrosis. Statistics were analyzed using an unpaired Student’s *t* test: statistical significance (p value) is indicated.

**Supplementary Figure S3.**
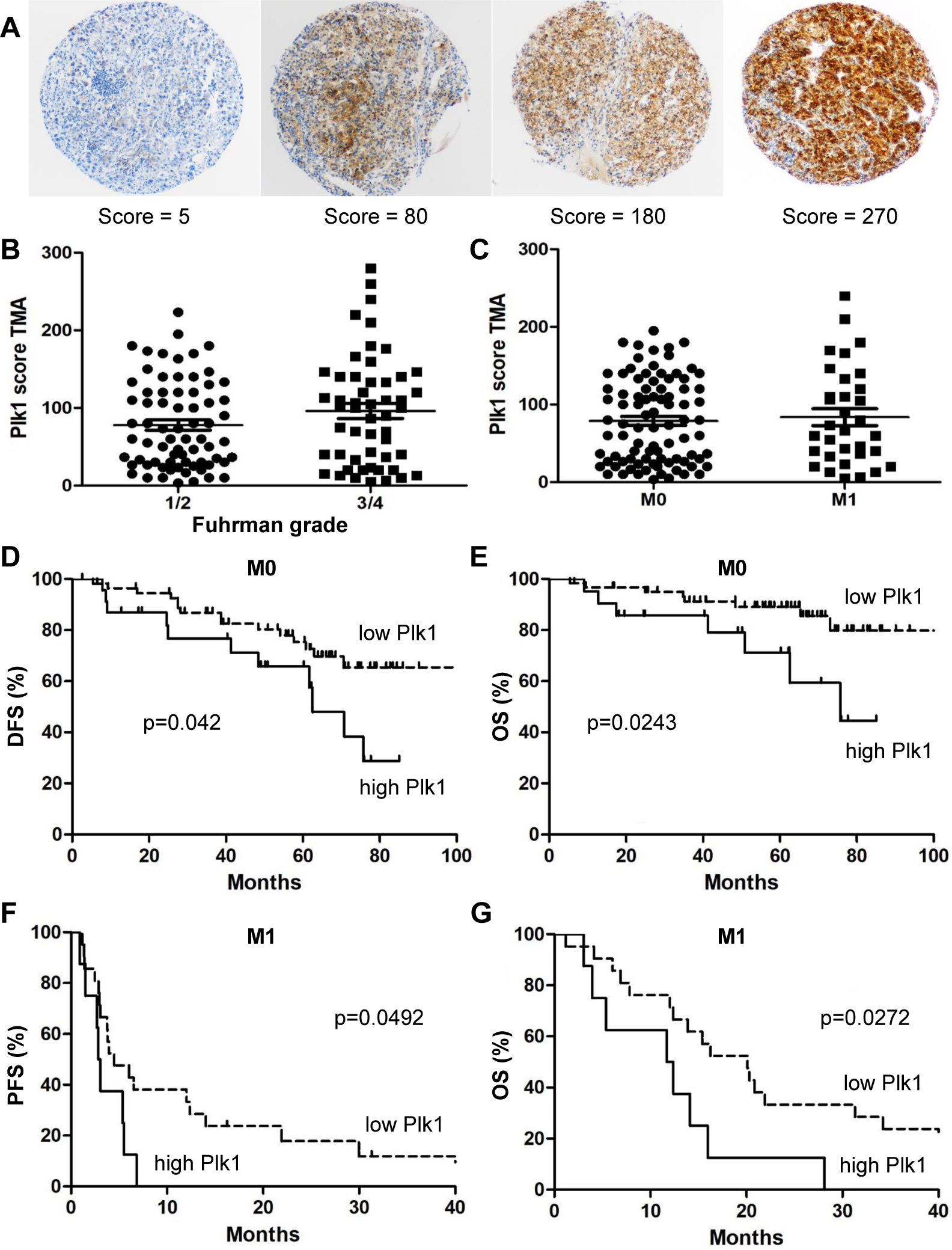
Detection of Plk1 on tumor sections by IHC. The percentage of cells and the intensity of the Plk1 labelling were evaluated. The Plk1 score was calculated form the percentage of labelled cells and the staining intensity. **A**, Representatives images are shown. **B**, The levels of the Plk1 score of ccRCC patients were compared to the Fuhrman grade 1/2 or 3/4. **C**, The levels of the Plk1 score of M0 ccRCC patients and of M1 ccRCC patients were compared. **D and E**, The levels of the Plk1 score of 101 M0 ccRCC patients correlated with DFS (**D**) or with OS (**E**). **F and G**, The levels of the Plk1 score of 30 M1 ccRCC patients correlated with PFS (**F**) or with OS (**G**). The third quartile value of the Plk1 score (120) was chosen as the reference. The Kaplan-Meier method was used to produce survival curves and analyses of censored data were performed using Cox models. Statistical significance (p values) is indicated (see Table S1).

**Supplementary Figure S4.**
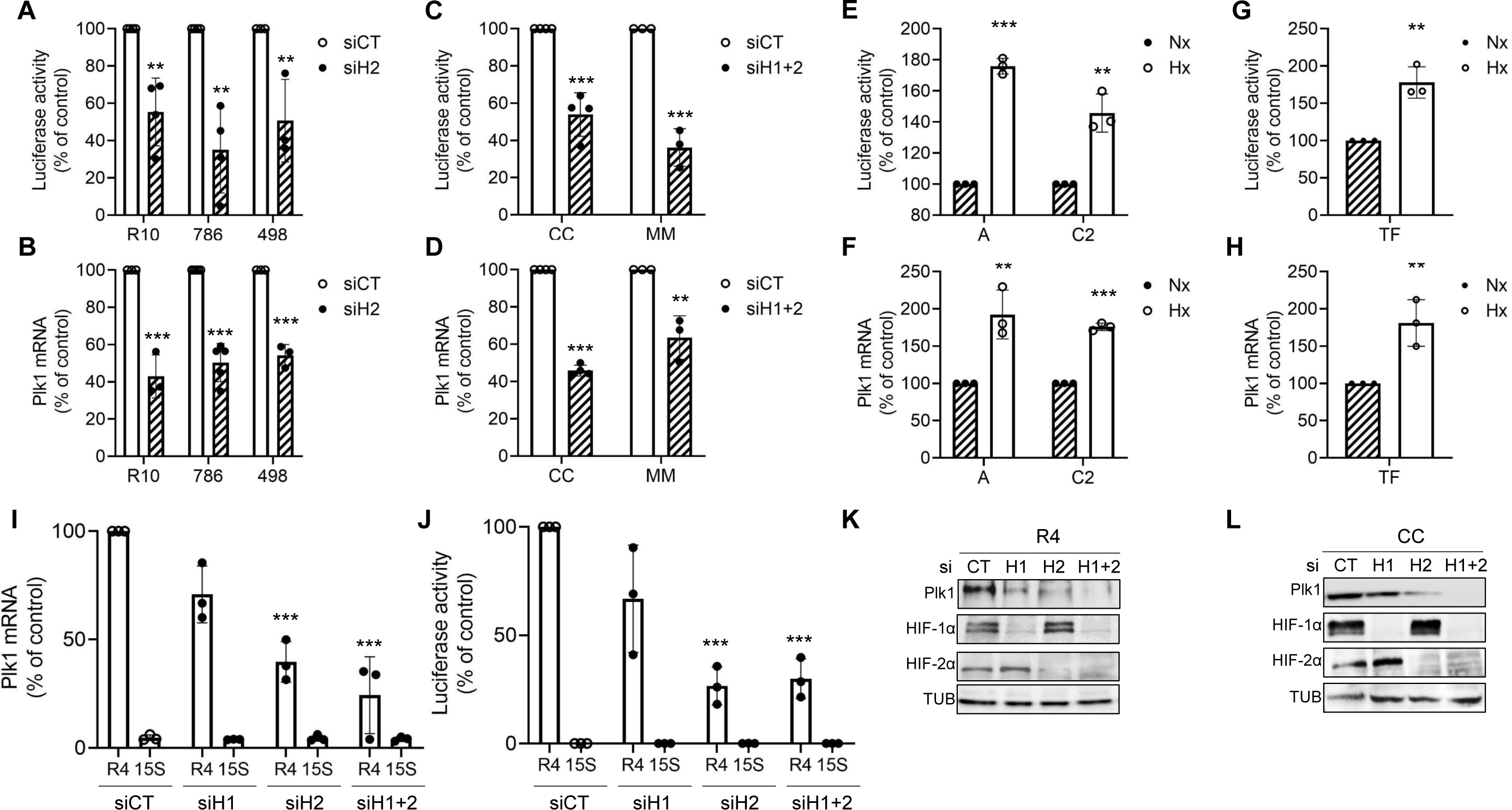
HIF-2 bound directly to the Plk1 promoter and regulated its transcription. **A to D**, ccRCC cell lines (VHL-inactivated) R10, 498, 786 (**A, B**), or primary ccRCC cells (VHL-inactivated) MM and CC (**C, D**) cells were transfected with siRNA against HIF-1α (H1), or HIF-2α (H2) or HIF-1α and HIF-2α (H1+2) for 24 h. Cells were then transfected with a renilla luciferase reporter gene under the control of the Plk1 promoter. The renilla luciferase activity normalized to the firefly luciferase (control vector) represented the readout of the Plk1 promoter activity (**A, C**). The Plk1 mRNA level was determined by qPCR (**B, D**). **E to H**, ccRCC cell lines (VHL-WT) ACHN and Caki2 (**E, F**), or primary ccRCC cells (VHL-WT) TF (**G, H**) were cultured in normoxia (Nx) or hypoxia 1% O2 (Hx) for 24 h. The renilla luciferase activity normalized to the firefly luciferase (control vector) represented the readout of the Plk1 promoter activity (**E, G**). The Plk1 mRNA level was determined by qPCR (**F, H**). **I and J**, ccRCC cell lines (VHL-inactivated) RCC4 (R4) or healthy renal cells (15S) were transfected with siRNA against HIF-1α (H1), or HIF-2α (H2) or HIF-1α and HIF-2α (H1+2). 48 hours after transfection, Plk1 mRNA levels were determined by qPCR (**I**). 24 hours later, cells were transfected with a renilla luciferase reporter gene under the control of the Plk1 promoter. The renilla luciferase activity, normalized to the firefly luciferase (control vector), was a readout of the *Plk1* promoter activity (**J**). **K and L**, ccRCC cell lines (VHL-inactivated) R4 (**K**) or primary ccRCC cells (VHL-inactivated) CC (**L**) were transfected with siRNA against HIF-2α (H2) for 48 h. Plk1, HIF-1α and HIF-2α expression was evaluated by immunoblotting. HSP90 served as a loading control. The graphs show the level of Plk1. The value of the control condition was considered as the reference value (100 %). Results are represented as the mean of three independent experiments ± SEM. Statistics were determined using an unpaired Student’s *t* test: * p<0.05, ** p<0.01, *** p<0.001.

**Supplementary Figure S5.**
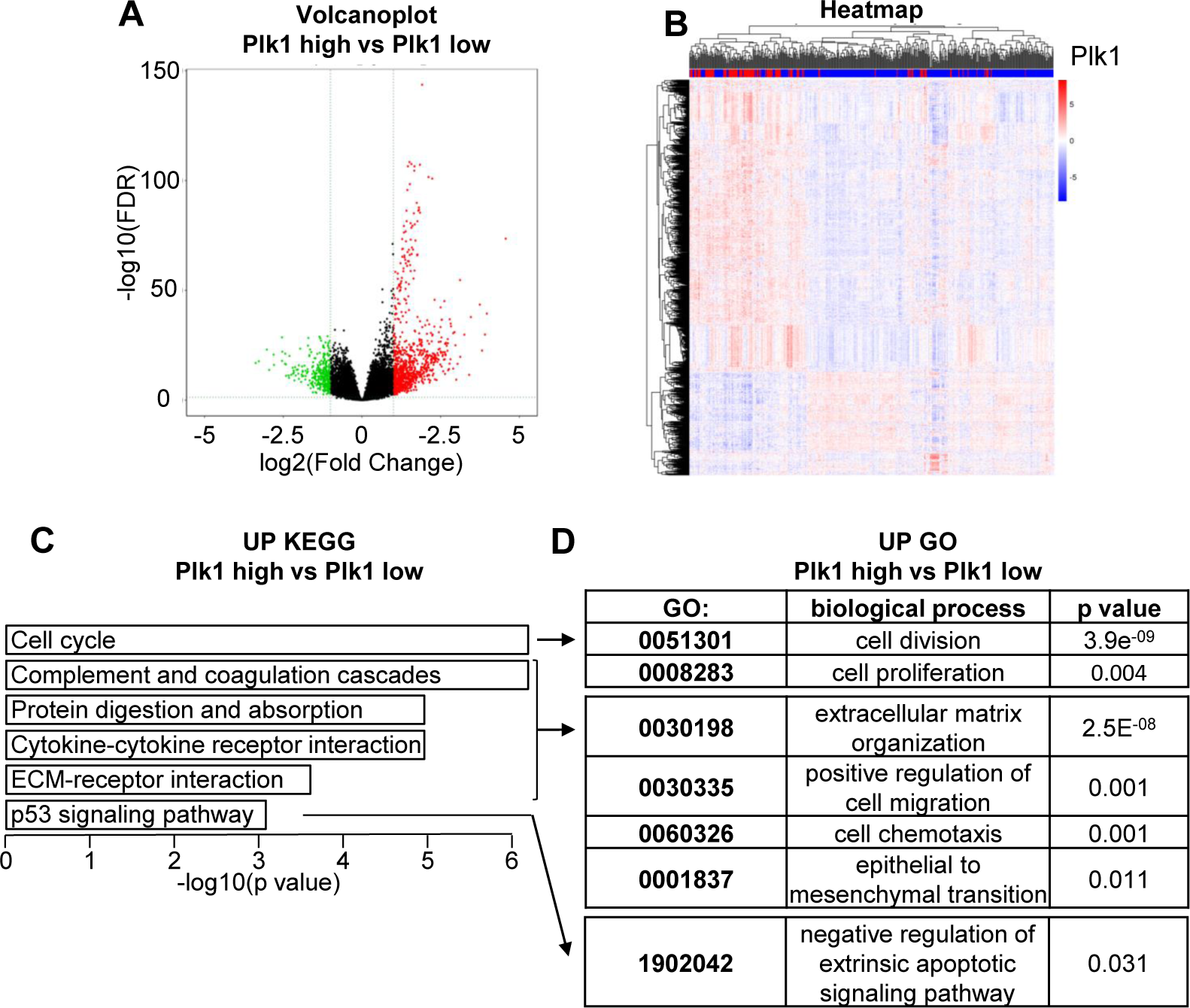
Analysis of pathway enrichment in ccRCC tumors according to the TCGA. **A**, Volcano plot showing the distribution of differentially expressed transcripts. 932 up-regulated genes in the Plk1 “high” group compared to the Plk1 “low” group are shown in red; 315 down-regulated genes are shown in green. Genes that were not differentially expressed (adj. p-value >0.05 and absolute log2(Fold change)>1) are shown in black. **B**, Heatmap comparing the normalized log2 expression (z score) of the differentially expressed genes between the 110 patients with high Plk1 expression and the 328 patients with low Plk1 expression to obtain differentially expressed genes. **C**, Graph of the top 6 enriched KEGG pathways from up-regulated genes. A Wilcoxon test was performed to obtain a p-value showing the differential significance. **D**, Graph of the enriched GO pathways (link KEGG pathway) from up-regulated genes. A Wilcoxon test was performed to obtain a p-value showing the differential significance.

**Supplementary Figure S6.**
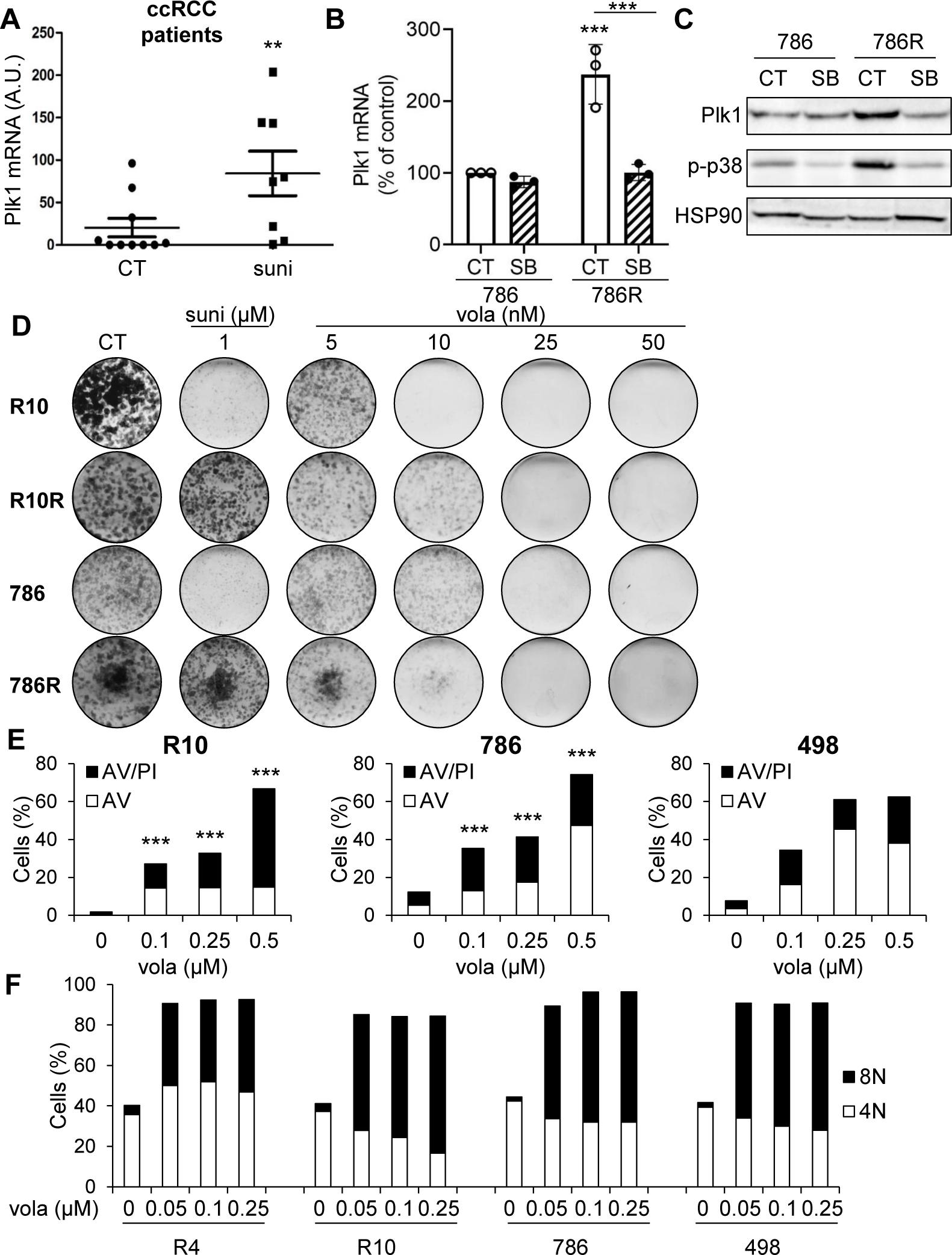
Sunitinib resistance are correlated with high Plk1 expression. **A**, The levels of Plk1 were determined by qPCR in tumors from patients either not treated or treated with sunitinib in a neoadjuvant setting (see Supplementary Table S4). **B and C**, 786 and 786R cells were treated with 20 µM SB203580 (p38 inhibitor, SB) for 48 h. The Plk1 mRNA level was obtained by qPCR (**B**). Plk1, p-p38, p38 expression were evaluated by immunoblotting. HSP90 served as a loading control (**C**). **D**, R10, R101R, 786 and 786R cells were incubated in the presence of volasertib (vola, 5 to 50 nM) or sunitinib (suni, 1 µM) and stained with Giemsa blue after 10 days. Results are representative of three independent experiments. **E and F**, ccRCC cells were treated with volasertib (vola) for 48 h. (**E**) Cell death was evaluated by flow cytometry. Cells were stained with the PI/annexinV (AV). Histograms show AV^+^/PI^−^ cells (apoptosis) and AV^+^/PI^+^ cells (post-apoptosis and/or others cell death). (**F**) Cells were labelled for 15 min with PI and analyzed by flow cytometry. Histograms represent the percentage of cells with a DNA content of 4N and 8N. Results are represented as means of three independent experiments ± SEM. Statistics were performed using an unpaired Student’s *t* test: ** p<0.01, *** p<0.001.

**Supplementary Figure S7.**
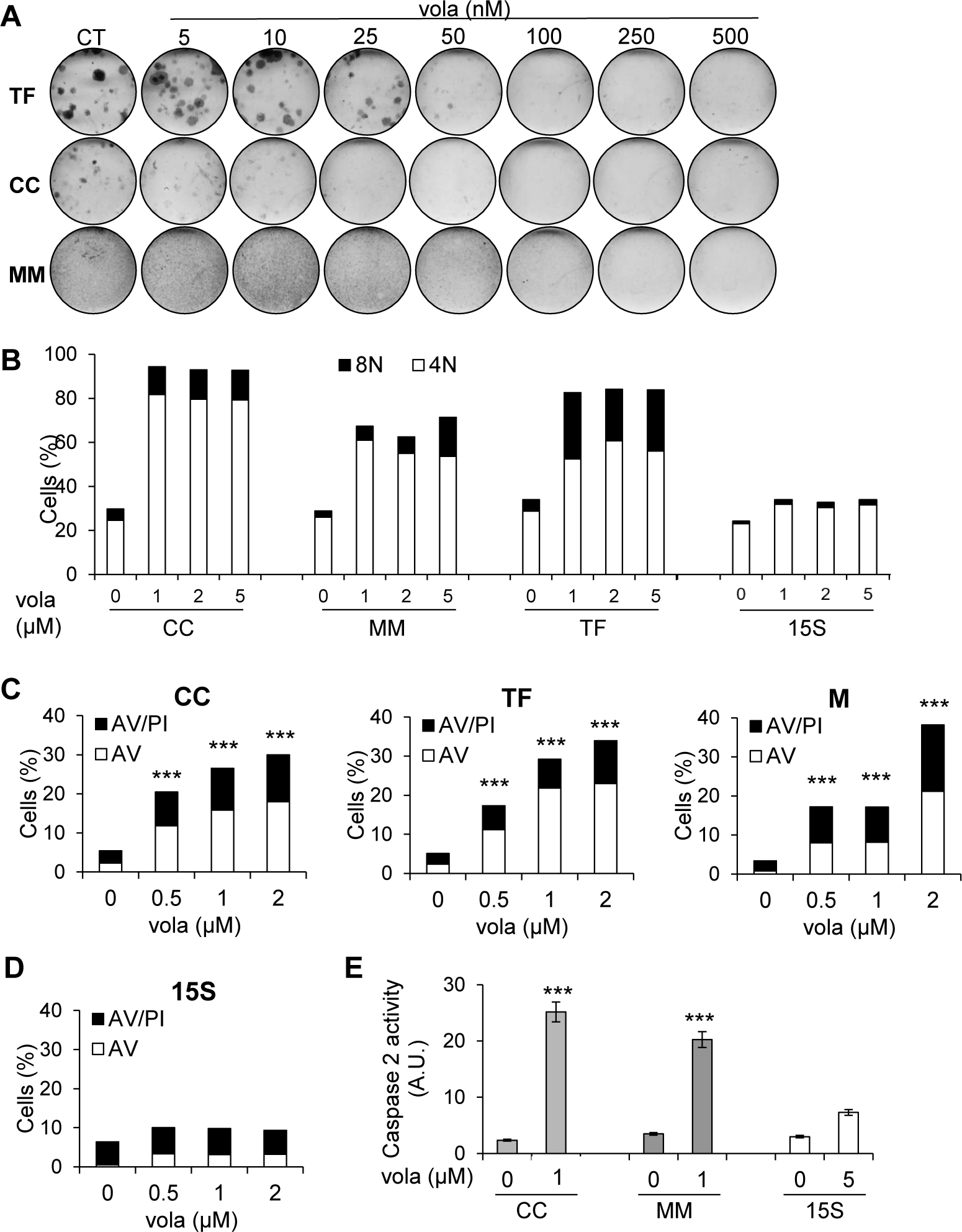
Volasertib induced polyploidy and apoptosis in ccRCC primary cells but not in normal cells. **A**, Primary ccRCC cells were incubated in the presence of volasertib (vola, 1 to 500 nM) and stained with Giemsa blue after 10 days. Results are representative of three independent experiments. **B**, Primary ccRCC cells and healthy renal cells were treated with volasertib (vola) for 48 h. Cells were labelled for 15 min with PI and analyzed by flow cytometry. Histograms represent the percentage of cells with a DNA content of 4N and 8N. Results are represented as means of three independent experiments. **C to E**, Primary ccRCC cells (**C, E**) and healthy renal cells (**D, E**) were treated with volasertib (vola) for 48h. Cell death was evaluated by flow cytometry. Cells were stained with the PI/ AV. Histograms showed AV^+^/PI^−^ cells (apoptosis) and AV^+^/PI^+^ cells (post-apoptosis and/or others cell death; **C, D**). The caspase 2 activity was evaluated using Ac-VDVAD-AMC as a substrate (**E**). Results are represented as means of three independent experiments ± SEM. Statistics were determined using an unpaired Student’s *t* test: * p<0.05, *** p<0.001.

**Supplementary Figure S8.**
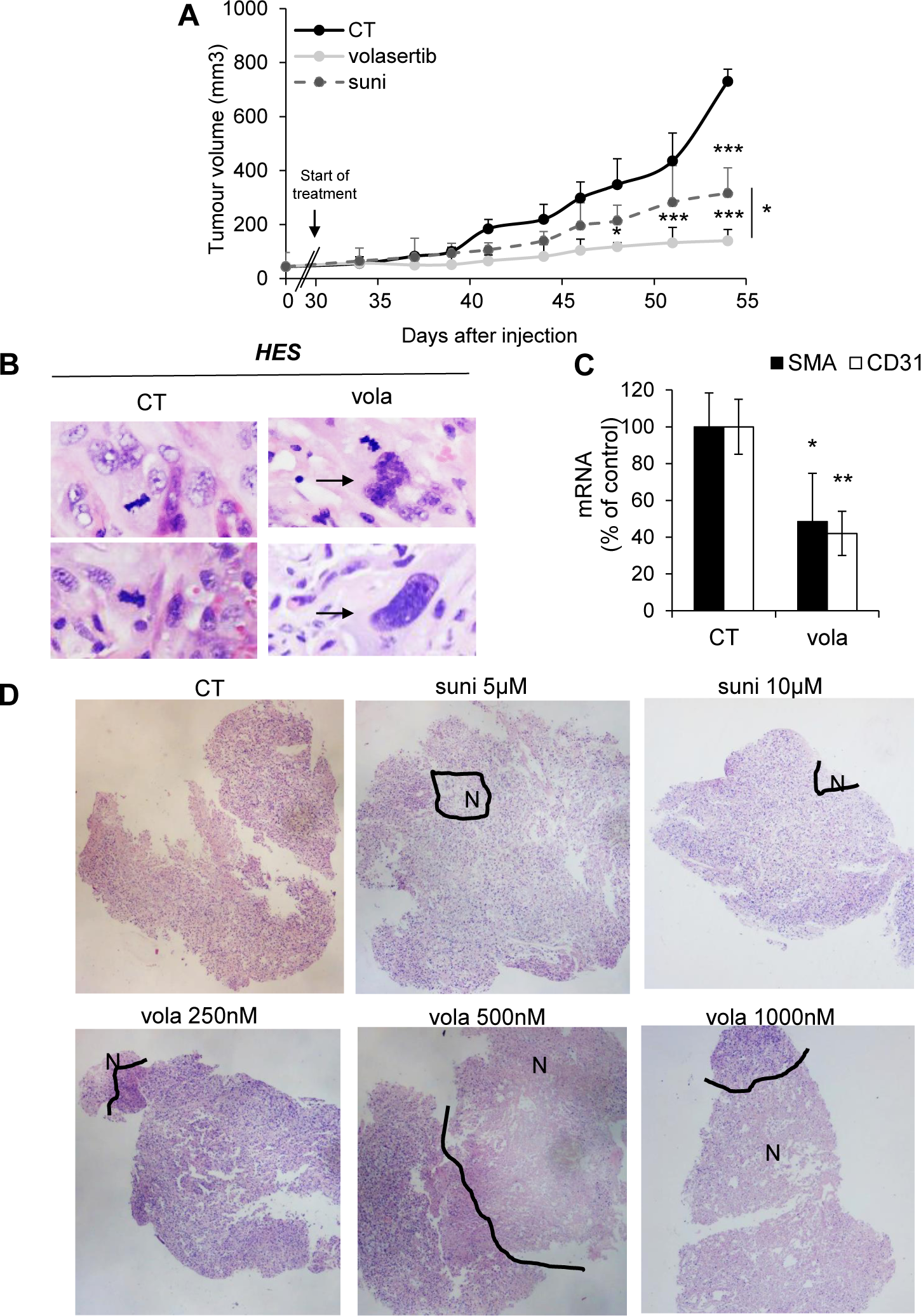
Volasertib inhibited the growth of experimental ccRCC more efficiently than sunitinib and induced necrosis in 3D ccRCC primary tumors. **A to C**, 7.10^6^ 786-O cells were subcutaneously injected into the flank of nude mice (*n*=8 per group). 30 days after injection, all mice developed tumors and were treated with a control solution or 20 mg/kg volasertib by gavage twice a week or 40 mg/kg sunitinib by gavage five times a week. **A**, The tumor volume was measured twice weekly as described in the materials and methods. **B**, HES coloration. Representative images are shown. Arrows indicate giant cells. **C**, The mRNA levels of CD31 and αSMA in tumors were determined by qPCR.One-way ANOVA was used for statistical comparison: * p<0.05, *** p<0.001. **D**, Representatives images of HES staining of 3D primary ccRCC tumors treated 72h with sunitinib (suni) or volasertib (vola). N: necrosis

**Supplementary Figure S9.**
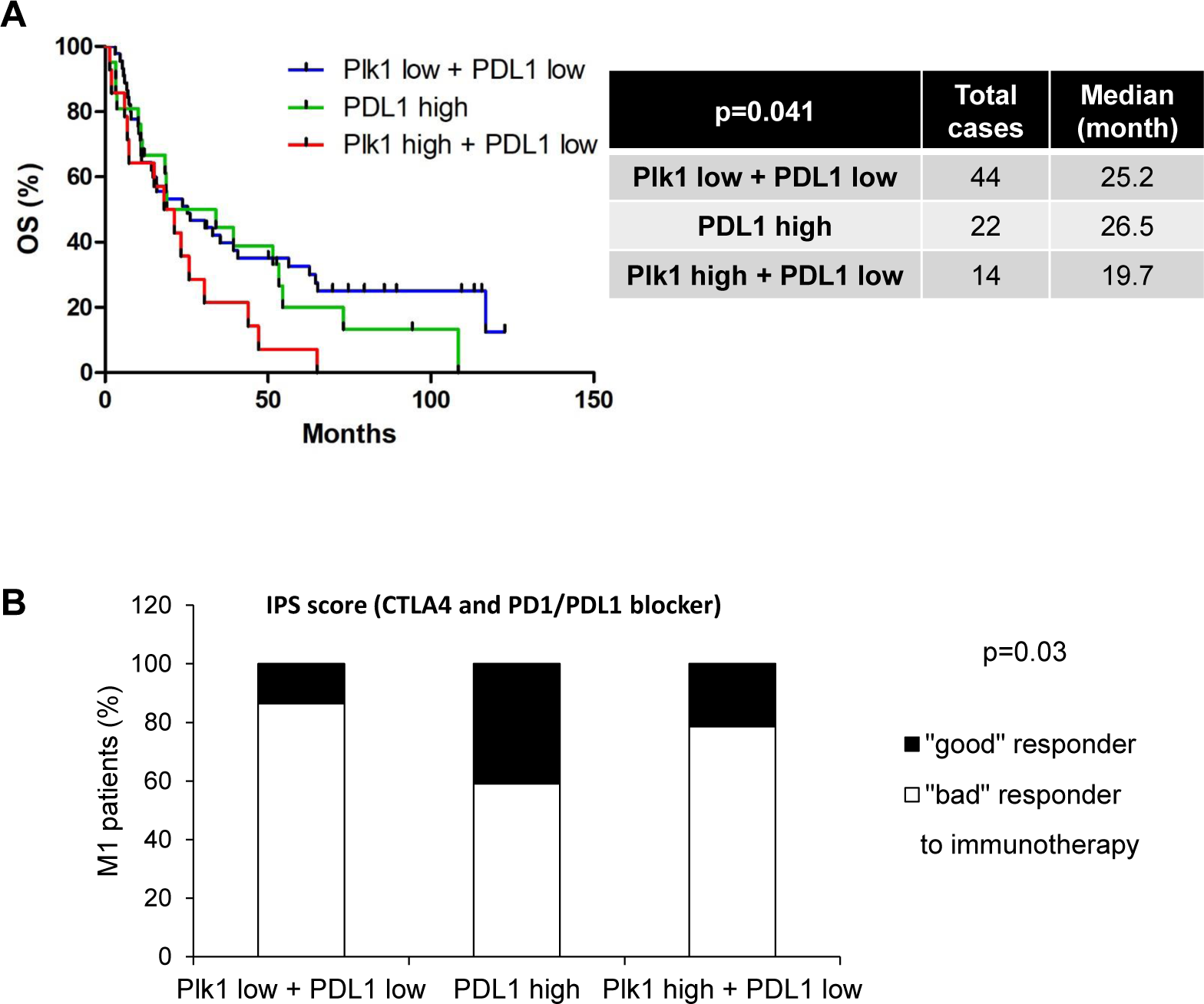
*In silico* analysis of TCGA and TCIA databases. These results are in whole or in part based upon data generated by the TCGA Research Network. **A**, Analysis of the cBioportal database, the levels of Plk1 and PDL1 mRNA in 80 metastatic (M1) ccRCC patients correlated with OS. **B**, Analysis of the TCIA database, the levels of Plk1 and PDL1 mRNA in 80 metastatic (M1) ccRCC patients correlated with the immunophenoscore (IPS, score predicting the response to CTLA4 and PD1/PDL1 immunotherapy). An IPS between 5 and 8 corresponded to “bad-intermediate” immunotherapy responder patients, and an IPS between 9 and 10 corresponded to “good” responder immunotherapy patients. **C**, The third quartile value of Plk1 and PDL1 expression was chosen as the reference. The Kaplan-Meier method was used to produce survival curves and analyses of censored data were performed using Cox models. Statistical significance (p values) is indicated.

**Supplementary Table S1.** High levels of Plk1 mRNA correlated to activation of the HIF pathway and to pejorative evolution of patients with different cancers. PFS and OS were calculated from patient subgroups in the cBioportal database (TCGA provisional cohort) with Plk1 mRNA levels that were less or greater than the third quartile. The total number of patients and the number of patients in each group are indicated. The levels of Plk1 mRNA (mRNA Expression z-Scores, RNA Seq V2 RSEM) are stratified into two groups expressing low or high levels (median cut-off) of Ca9, Oct4 or Glut1. Ca9 served as a marker of HIF-1 activity, Oct4 served as a marker of HIF-2 activity and Glut1 served as a marker of HIF-1 and HIF-2 activities. Four groups related to Plk1 expression were defined: **A**, HIF-1 and HIF-2 dependency (Ca9, Oct4 and Glut1); **B**, HIF-1 dependency (Ca9 and Glut1); **C**, HIF-2 dependency (Oct4 and Glut1) and **D**, HIF-1 or HIF-2 independency (none of them). The different cancers analyzed were the following: Kidney Renal Clear Cell Carcinoma (ccRCC), Kidney Renal Papillary Cell Carcinoma (pRCC), Kidney Chromophobe (chRCC), Breast Invasive Carcinoma (Breast), Head and Neck Squamous Cell Carcinoma (HNSCC), Liver Hepatocellular Carcinoma (liver), Lung Adenocarcinoma (Lung), Skin Cutaneous Melanoma (melanoma), Pancreatic Adenocarcinoma (pancreatic), Sarcoma, Uterine Corpus Endometrial Carcinoma (uterine). Statistical significance (p value) is indicated.

| Breast (960 patients) |  |  |  |
| --- | --- | --- | --- |
|  | low Plk1 | high Plk1 | p value |
| PFS (months) | >160 | 160 | 0.003 |
| OS (months) | 128 | 130 | 0.014 |
|  | high Ca9 | high Oct4 | high Glut1 |
| Plk1 mRNA | + | + | + |
| p value | <0.001 | <0.001 | <0.001 |

| Liver (373 patients) |  |  |  |
| --- | --- | --- | --- |
|  | low Plk1 | high Plk1 | p value |
| PFS (months) | 27 | 11 | 3e-4 |
| OS (months) | 70 | 30 | 1e-4 |
|  | high Ca9 | high Oct4 | high Glut1 |
| Plk1 mRNA | + | + | + |
| p value | <0.001 | <0.001 | <0.001 |

| Sarcoma (263 patients) |  |  |  |
| --- | --- | --- | --- |
|  | low Plk1 | high Plk1 | p value |
| PFS (months) | 43 | 18 | 0.004 |
| OS (months) | 80 | 49 | 0.011 |
|  | high Ca9 | high Oct4 | high Glut1 |
| Plk1 mRNA | + | + | + |
| p value | 0.006 | <0.001 | <0.001 |

**B****HIF-1 dependency**
| pRCC (413 patients) |  |  |  |
| --- | --- | --- | --- |
|  | low Plk1 | high Plk1 | p value |
| PFS (months) | 104 | 26 | 2e-9 |
| OS (months) | >160 | 58 | 8e-10 |
|  | high Ca9 | high Oct4 | high Glut1 |
| Plk1 mRNA | + | -/+ | + |
| p value | 0.001 | ns | <0.001 |

| chrRCC (66 patients) |  |  |  |
| --- | --- | --- | --- |
|  | low Plk1 | high Plk1 | p value |
| PFS (months) | >160 | 160 | 0.003 |
| OS (months) | >160 | >160 | 0.003 |
|  | high Ca9 | high Oct4 | high Glut1 |
| Plk1 mRNA | + | -/+ | + |
| p value | 0.063 | ns | 0.013 |

| HNSCC (552 patients) |  |  |  |
| --- | --- | --- | --- |
|  | low Plk1 | high Plk1 | p value |
| PFS (months) | 76 | 4 | 0.056 |
| OS (months) | 65 | 32 | 0.003 |
|  | high Ca9 | high Oct4 | high Glut1 |
| Plk1 mRNA | + | -/+ | + |
| p value | <0.001 | ns | ns |

| Lung (230 patients) |  |  |  |
| --- | --- | --- | --- |
|  | low Plk1 | high Plk1 | p value |
| PFS (months) | 40 | 23 | 0.053 |
| OS (months) | 50 | 36 | 0.018 |
|  | high Ca9 | high Oct4 | high Glut1 |
| Plk1 mRNA | + | -/+ | + |
| p value | 0.05 | ns | <0.001 |

| Melanoma (472 patients) |  |  |  |
| --- | --- | --- | --- |
|  | low Plk1 | high Plk1 | p value |
| PFS (months) | 55 | 48 | ns |
| OS (months) | 102 | 50 | 0.003 |
|  | high Ca9 | high Oct4 | high Glut1 |
| Plk1 mRNA | + | -/+ | + |
| p value | <0.001 | ns | <0.001 |

| Pancreatic (179 patients) |  |  |  |
| --- | --- | --- | --- |
|  | low Plk1 | high Plk1 | p value |
| PFS (months) | 12 | 8 | 0.001 |
| OS (months) | 21 | 15 | 0.001 |
|  | high Ca9 | high Oct4 | high Glut1 |
| Plk1 mRNA | + | -/+ | + |
| p value | <0.001 | ns | <0.001 |

**C****HIF-2 dependency**
| ccRCC (413 patients) |  |  |  |
| --- | --- | --- | --- |
|  | low Plk1 | high Plk1 | p value |
| PFS (months) | 124 | 33 | 3e-9 |
| OS (months) | >160 | 43 | 2e-12 |
|  | high Ca9 | high Oct4 | high Glut1 |
| Plk1 mRNA | -/+ | + | + |
| p value | ns | 0.007 | <0.001 |

**D HIF-1 or HIF-2 independency**
| Uterine (177 patients) |  |  |  |
| --- | --- | --- | --- |
|  | low Plk1 | high Plk1 | p value |
| PFS (months) | >160 | > 160 | 0.047 |
| OS (months) | >160 | 102 | 0.007 |
|  | high Ca9 | high Oct4 | high Glut1 |
| Plk1 mRNA | -/+ | -/+ | -/+ |
| p value | ns | ns | ns |

**Supplementary Table S2.**
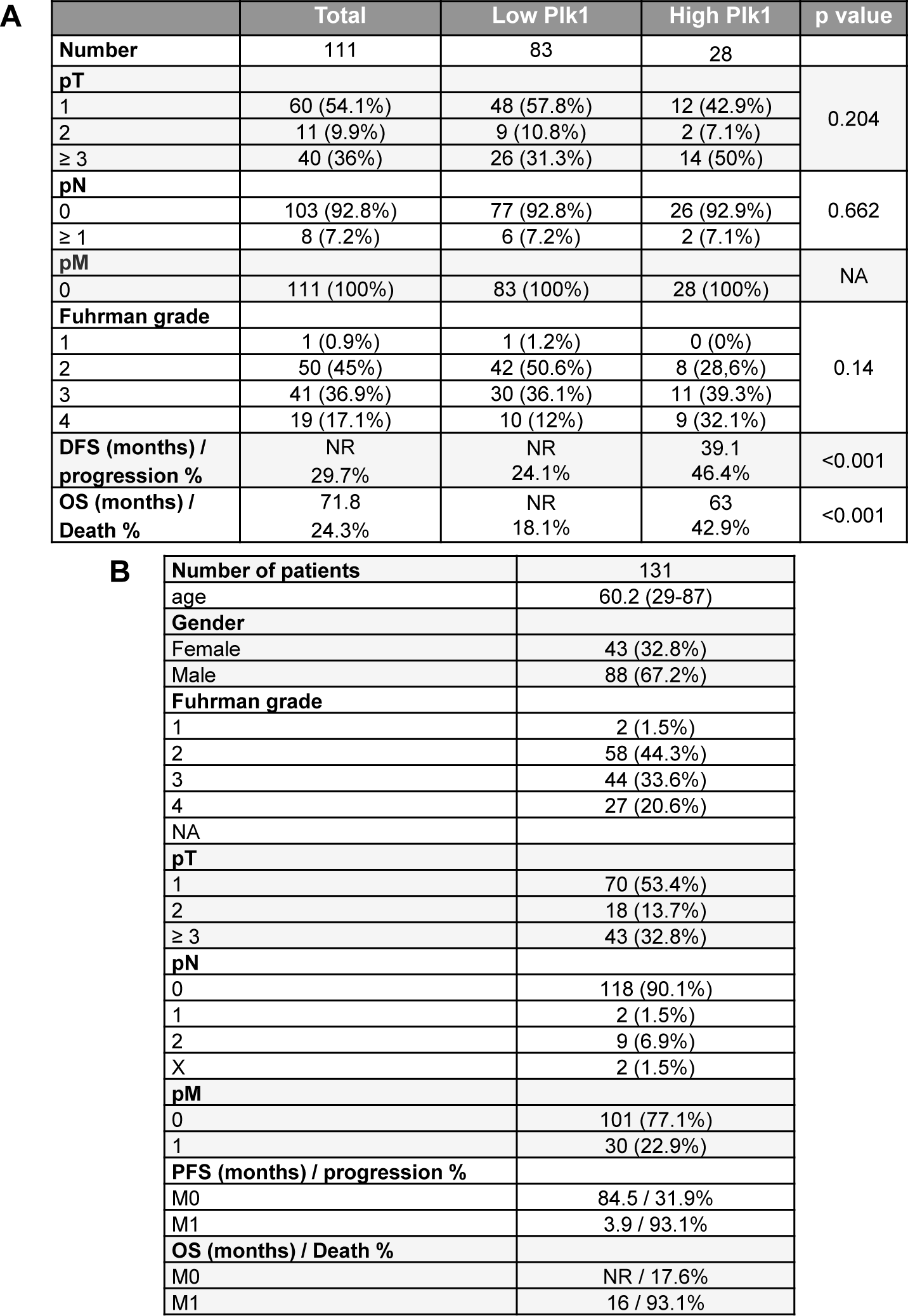
The characteristics of the M0 patients included in the study and a multivariate analysis. **A**, Patient characteristics and univariate analysis with the Fisher or Ki^2^ test. Statistical significance (p values) is indicated (see Figure 1). **B**, Patient characteristics, included in Supplemntary Fig. S3.

**A**
|  | Total | Low Plk1 | High Plk1 | p value |
| --- | --- | --- | --- | --- |
| **Number** | 111 | 83 | 28 |  |
| **pT** |  |  |  | 0.204 |
| 1 | 60 (54.1%) | 48 (57.8%) | 12 (42.9%) |  |
| 2 | 11 (9.9%) | 9 (10.8%) | 2 (7.1%) |  |
| ≥ 3 | 40 (36%) | 26 (31.3%) | 14 (50%) |  |
| **pN** |  |  |  | 0.662 |
| 0 | 103 (92.8%) | 77 (92.8%) | 26 (92.9%) |  |
| ≥ 1 | 8 (7.2%) | 6 (7.2%) | 2 (7.1%) |  |
| **pM** |  |  |  | NA |
| 0 | 111 (100%) | 83 (100%) | 28 (100%) |  |
| **Fuhrman grade** |  |  |  | 0.14 |
| 1 | 1 (0.9%) | 1 (1.2%) | 0 (0%) |  |
| 2 | 50 (45%) | 42 (50.6%) | 8 (28,6%) |  |
| 3 | 41 (36.9%) | 30 (36.1%) | 11 (39.3%) |  |
| 4 | 19 (17.1%) | 10 (12%) | 9 (32.1%) |  |
| **DFS (months) / progression %** | NR<br>29.7% | NR<br>24.1% | 39.1<br>46.4% | <0.001 |
| **OS (months) / Death %** | 71.8<br>24.3% | NR<br>18.1% | 63<br>42.9% | <0.001 |

**B**
|  |  |
| --- | --- |
| **Number of patients** | 131 |
| age | 60.2 (29-87) |
| **Gender** |  |
| Female | 43 (32.8%) |
| Male | 88 (67.2%) |
| **Fuhrman grade** |  |
| 1 | 2 (1.5%) |
| 2 | 58 (44.3%) |
| 3 | 44 (33.6%) |
| 4 | 27 (20.6%) |
| NA |  |
| **pT** |  |
| 1 | 70 (53.4%) |
| 2 | 18 (13.7%) |
| ≥ 3 | 43 (32.8%) |
| **pN** |  |
| 0 | 118 (90.1%) |
| 1 | 2 (1.5%) |
| 2 | 9 (6.9%) |
| X | 2 (1.5%) |
| **pM** |  |
| 0 | 101 (77.1%) |
| 1 | 30 (22.9%) |
| **PFS (months) / progression %** |  |
| M0 | 84.5 / 31.9% |
| M1 | 3.9 / 93.1% |
| **OS (months) / Death %** |  |
| M0 | NR / 17.6% |
| M1 | 16 / 93.1% |

**Supplementary Table S3.** The IC50 of the different Plk1 inhibitors: volasertib, BI2536, ON01910, GSK461364, in different ccRCC cell lines, primary ccRCC cells and normal kidney cells. Cells were treated with volasertib for 48 h. Cell viability was measured with XTT assays and the IC50 was determined.

| Type of cells | Cell name's | IC50 Volasertib (nM) | IC50 BI2536 (nM) | IC50 ON01910 (nM) | IC50 GSK461364 (nM) |
| --- | --- | --- | --- | --- | --- |
| ccRCC cell line | R4 | 55<br>+/- 6 | 8<br>+/- 2 | 20<br>+/- 5 | 25<br>+/- 3 |
|  | R10 | 85<br>+/- 10 | 7<br>+/- 2 | 20<br>+/- 4 | 23<br>+/- 4 |
|  | 786 | 80<br>+/- 10 | 25<br>+/- 3 | 10<br>+/- 2 | 50<br>+/- 5 |
|  | 498 | 90<br>+/- 8 | 30<br>+/- 4 | 48<br>+/- 3 | 40<br>+/- 4 |
| Primary ccRCC cells | CC | 260<br>+/- 13 | 50<br>+/- 4 | 25<br>+/- 9 | 80<br>+/- 24 |
|  | TF | 750<br>+/- 14 | 150<br>+/- 15 | 80<br>+/- 12 | 200<br>+/- 21 |
|  | M | 620<br>+/- 25 | 120<br>+/- 12 | 70<br>+/- 6 | 200<br>+/- 14 |
| Normal kidney cells | 14S | 3 300<br>+/- 100 | >500 | 90<br>+/- 17 | >500 |
|  | 15S | 2 500<br>+/- 111 | >500 | 95<br>+/-13 | >500 |
|  | 18S | 2 400<br>+/- 89 | >500 | 100<br>+/-9 | >500 |

## Supplementary Materials and methods

### Colony formation assay

ccRCC cells (500 cells per condition) were treated or not with sunitinib or volasertib. Colonies were detected after 10 days of culture. Cells were then washed, fixed and stained with GEMSA (Sigma).

### Patient TMA

Tissue microarrays (TMA) of primary tumor samples of ccRCC patients were obtained from the Bordeaux University Hospital. The Plk1 score was calculated from the percentage of labelled cells (0 to 100%) multiplied by the staining intensity (0 to 3). The DFS, PFS and OS were calculated from patient subgroups with Plk1 expression that was less or greater than the third quartile (score = 120) IHC score (Fig. S3 and Table S1).

### Immunohistochemistry TMA

Samples were collected with the approval of the Local Ethics committee. Sections from blocks of formol-fixed and paraffin-embedded tissue were examined for immunostaining for Plk1. After deparaffinization, hydration and heat-induced antigen retrieval, the tissue sections were incubated for 20 min at room temperature with monoclonal anti-Plk1 antibody (Abcam, ab109777) diluted at 1:100. Biotinylated secondary antibody (DAKO) was applied and binding was detected with the substrate diaminobenzidine against a hematoxylin counterstain.

### Neoadjuvant patients for qPCR analysis

Samples (tumor sections) were obtained from Nice, Bordeaux and Monaco hospitals. The patients’ characteristics have already been described (1). Patients were treated for at least two months before surgery (Fig. S6*A*).

### Gene expression microarray analysis

Normalised RNA sequencing (RNA-Seq) data produced by The Cancer Genome Atlas (TCGA) were downloaded from cBioportal (www.cbioportal.org, TCGA Provisional; RNA-Seq V2). Data were available for 503 of the 536 ccRCC tumor samples TCGA subjected to mRNA expression profiling. The subtype classifications were obtained through cBioPortal for Cancer Genomics and the 33 samples lacking classification were discarded. The non-metastatic group contained 424 patients and the metastatic group contained 79 patients. The results published here are in whole or in part based upon data generated by the TCGA Research Network: http://cancergenome.nih.gov/(2,3) The Kaplan-Meier method was used to produce overall survival curves. The effect of Plk1 and its odds-ratio was estimated using a Cox model adjusted to the expression of other genes and important patient characteristics.

To evaluate the effect of Plk1 gene expression on ccRCC, we used the Kidney Renal Clear Cell Carcinoma (KIRC) dataset from The Cancer Genome Atlas (TCGA) (4). RNA-seq and clinical data were downloaded from the TCGA data portal (https://portal.gdc.cancer.gov). RNA-seq data were normalised using the Bioconductor package DESeq2 and log2 transformed. The patients were separated into two groups with either a high or low Plk1 expression level (third quartile cut off). We then performed a differential analysis between the two groups of patients.

P values were adjusted for multiple testing using the Benjamini and Hochberg procedure, which controls the false discovery rate (FDR).

We then performed a functional and pathways enrichment analysis on differentially expressed genes (FDR < 0.05 and absolute log2(Fold Change) > 1) based on KEGG, Gene Ontology and Reactome databases using the geneSCF tool (4). The terms are considered significant only if enriched with a p value < 0.05.

